# Hospital preparedness in epidemics by using simulation. The case of COVID-19

**DOI:** 10.1101/2020.08.12.20173328

**Authors:** Daniel Garcia-Vicuña, Laida Esparza, Fermin Mallor

## Abstract

This paper presents a discrete event simulation model to support the decision-making concerned with the short-term planning of the necessary hospital resources, especially Intensive Care Unit (ICU) beds, to face outbreaks, as the SARS-CoV-2. Being used as a short-term forecasting tool, the simulation model requires an accurate representation of the current system state and high fidelity in mimicking the system dynamics from that state. The two main components of the simulation model are the stochastic modeling of the admission of new patients and the patient flow through the hospital facilities. For the patient arrival process, we analyze different models based on growth curves of the twenty most affected countries (until June 15) and propose the use of the Gompertz curve. The length of stay is divided into several stages, each one modeled separately. We analyze the starting of the simulation model, which requires different procedures depending on the information available about the patients currently hospitalized. We also report the use of this simulation model during the COVID-19 outbreak in the Autonomous Community of Navarre, in Spain. Every day, the research team informed the regional logistic team in charge of planning the health resources, who programmed the ward and ICU beds based on the resulting predictions.

## 1 Introduction

COVID-19 pandemic presents an important threat to global health. Since the outbreak in China in early December 2019, the number of patients confirmed to have the disease has exceeded seventeen million, and more than 650,000 people have died from COVID-19 infection (up to 30 July 2020 https://coronavirus.jhu.edu/map.html). Regularly updated information on the COVID-19 outbreak is also available on the European Centre for Disease Prevention and Control’s (ECDC) website [1], the European Commission’s (EC) website [2], and the World Health Organization’s (WHO) website [3]. This outbreak has changed how health care is delivered, and it has affected the operation in hospitals, which have experienced an increase in demand. The pandemic has impacted on the Intensive Care Units (ICU), which involve highly specialized personnel and expensive technical sanitary material, that require time to their planning and acquisition. Therefore, an accurate prognosis of the necessary resources is needed to efficiently manage the scarce resources and to provide the best possible care to patients. The precision of the predictions allows preparing the response and helping to save lives. Usually, the hospitalization bed is still widely used as a hospital (ICU) management parameter both at the strategic and operational levels.

Hospitals are complex systems evolving in a stochastic environment whose uncertainty is even higher in pandemic periods because of the lack of knowledge about the spread of the disease and its consequences on patients. In this unsettle context, simulation emerges as a suitable analytical tool, since it can represent the complexity of the analyzed system and the variability and uncertainty of its environment. Besides, simulation can be used in combination with other analytical techniques. The literature contains numerous bibliographical references relating to the use of simulation models for decision making in the healthcare context, as it is exposed in Section 2. Most of the applications use the simulation to support strategic decisions, usually for dimensioning of resources, for their scheduling, or for their management. All these cases require the design of a simulation model to reproduce the performance of the health system in its stationary state and evaluate the resource levels, the patient flow management policies, and the decision making process in the long term. The recommendations obtained from the simulation analysis are meant to be implemented in the health system and last for a certain time horizon.

However, a simulation model designed to help to make tactical decisions related to the provision of specialized health resources during the current outbreak has to focus on the transition period, to project in the future the current state of the hospital. A discrete event simulation (DES) model is presented in this research that combines a dynamic forecast method to predict (simulate) the new incoming patients and the reproduction of the patient flow through the health system. The simulation output provides scenarios of future utilization of resources that keep informed the health authorities of future needs and give them time for their planning. Therefore, the main feature of the simulation model presented here is its capacity of reproducing the evolution of the health system from its current state to be used as a forecasting tool. The correct representation of the current state of the hospital and the methodology used to start the simulation plays a crucial role in the accuracy of the simulation results. Nevertheless, these two aspects strongly depend on the available information about the hospital and its admitted patients. In some cases, there could be data to the patient level, that is, with knowledge of the admission and discharge dates (including transferring dates between facilities or departments inside the hospital) while in other cases only aggregated information is available, for example, the total number of admitted patients each day. The simulation model we have developed is flexible enough to handle the different levels of information.

The main contribution of this paper is the proposal of a new simulation framework to enable the prediction in the short term (from days to one month) the need for critical resources to provide healthcare to COVID-19 patients. The simulation framework can be adapted to be applied to other future outbreaks. To reach this main contribution our research includes:

- A simulation method of patient arrivals based on population growth (PG) models.
- A statistical analysis of four different PG models to analyze their accuracy to represent the expansion of the pandemic and their forecasting capacity.
- The representation of the current state of the health system and the beginning of the simulation from this state by considering four different information levels for the admitted patients.
- A dynamic statistical analysis of the patient flow through the health facilities.
- The combination of all elements in a DES model flexible enough to recreate scenarios based on stochastic models fitted to data (data-driven prediction), scenarios defined by expert judgment, and a mixture of both.
- From a practical point of view, this paper also reports a successful real application of simulation to support an important decision-making process essential for the health of patients in an Autonomous Community of Spain (Navarre).

The rest of the paper is organized as follows. Section 2 presents a literature review of papers dealing with the use of quantitative methods to predict needs in health care systems and improve their management. Section 3 studies the adequacy of PG models to predict the evolution of the spread of a pandemic. The modeling of the patient flow through the hospital is presented in Section 4. The structure of the DES model and the methodology to begin the simulation under different available data are included in Section 5. Section 6 describes the simulator, with its inputs and outputs. Results of the application of the simulation model to the Autonomous Community of Navarre (Spain) are included in Section 7. Finally, Section 8 ends the paper with the conclusions of this work.

## 2 Related literature

Simulation is one of the most suitable analytical tools for the analysis of complex systems, as the health systems are, which is reflected in numerous articles in the specialist literature describing the use of simulation models for decision-making in the healthcare context. DES has been applied to model and analyze all aspects of logistics management in healthcare. In particular, it has been used to improve patient flow management, bed-planning, waiting list management, health service design, medical staff scheduling, etc. For reviews of the use of simulation models in healthcare, we refer to [4–7]. Usually, these simulation models focus on studying the stationary state of the health system to support strategic decisions related to the dimensioning of resources or designing management policies.

The ultimate goal of these models is to reconcile resource availability with demand in order to provide high-quality healthcare to patients while maintaining a reasonable level of human and technological resources. Problems analyzed into this framework are patient flow [8, 9], bed planning [10–12], health service design [13] and medical staff scheduling [14], among others. Although discrepancies between assumptions made in mathematical simulation models and behavior of real health systems reported in the medical literature have been pointed out [15], there is no doubt about the usefulness of simulation models for the analysis of relevant problems in complex health systems.

However, simulation not only helps to ensure that medical staff and facilities are offering the highest quality services but also increases the likelihood of following best practices. Since the pandemic began, all national governments and the World Health Organization have extensively used simulation modelling to decide the best strategies to reduce the impact of COVID-19.Currie et al. [16] identify challenges from this disease and discuss how simulation modelling could help to support decision-makers in making the most informed decisions.

The accuracy of a simulation model to predict the necessary resources during a pandemic depends on an accurate model to predict the arrival of new patients to the health service. Infectious disease prediction models mainly include differential equation prediction models based on population dynamics [17, 18]. These mathematical models are essential to understand the course of the epidemic and to plan effective control strategies [19–21]. One of the most widely used models in the human-to-human transmission is the SIR model [22]. The individuals of the population are divided into different categories, each one considered as a possible state for the individual: S (Susceptible), I (Infected), and R (Remove). The population in each state is calculated over time from the estimation of the transition rates among these states. By increasing the complexity of this model, it is possible to recreate the spread of specific epidemics. In fact, for the COVID-19 pandemic, extensions of the classical SIR model have been developed [23–27], as well as stochastic transmission models [28, 29]. However, this kind of models are complicated and need strong assumptions and simplifications, because they are based on a set of few differential equations with initial conditions and a number of adaptive parameters [30–33].

Therefore, growth population models suppose a simpler alternative to model the accumulated number of infected people. Growth curves are found in a wide range of areas, such as fishery research [34, 35], biology [36] or other infectious disease outbreaks [37–40]. Specifically, Logistic, Gompertz, Rosenzweig, and Richards models have been already used to model the spread of outbreaks such as A/H1N1 and Ebola in [41]. In relation to COVID-19 disease, several papers have been found in the literature which develop a growth model to predict new positive cases in different countries such as China [42], India [43], Spain [44], and other European countries [45]. These mathematical models present a set of mathematical equations that include some adaptive parameters that can be determined numerically based on available real data [46]. So, the model can be used daily (by updating the number of positive cases) and automatically adapt to the evolution of each parameter.

If all the mathematical models mentioned in the previous paragraphs are capable of fitting well to the real data, it would be possible to predict quite accurately what might happen in the future (e.g., emergency planning, resource allocation) [47–49]. This is very important especially for those resources that are normally scarce in a hospital, such as ICU beds. Manca et al. [50] present and discuss a few regression models developed using historical data of ICU patients and deaths during COVID-19 pandemic. They are capable of reproducing the bed occupancy curve using mathematical models, which can be very useful from the point of view of decision-making and emergency planning in future pandemics.

Besides, in recent decades, simulation in healthcare has become a new way of learning through experiences with advanced technology [51, 52] With this modern education and training technique, healthcare professionals can learn new cognitive, technical, and behavioral skills. Before working in real-world scenarios, where real patients are treated, both professionals and students can use this experimental learning style to develop their skills and expand knowledge without taking any risk in their decision-making processes [53]. Simulation models, as the one presented in this paper, can serve also for learning the management of health care services in emergencies. One of the most critical decisions in periods of scarcity of resources is their assignment to patients, especially when these decisions can make the difference between recovery and death, as it could happen with the admission of patients to ICUs. This triage is difficult to implement during pandemics where the situations of a scarcity of resources are further aggravated. Different protocols for ICU triage during a pandemic have been suggested in [54–56]. Foreseeing the increasing need for beds is essential to avoid ethical dilemmas [15, 57]. According to Utley et al. [58], “the impact of triage is dependent on the level of demand and on the scale of achievable differences between included and excluded groups in terms of anticipated length of stay and critical care survival”. A simulation model can help better planning of critical resources, during a pandemic; and as an off-line tool, it can be used as a learning tool to test new protocols for triage, because sometimes triage is not as effective as might be expected, and other hard-to-anticipate factors must be considered.

## 3 Modelling the patient arrival pattern

In this section, the adequacy of population models to predict new cases is studied. First, four different models are compared statistically to select the more suitable model. Then, we described the simulation of patient arrival to the hospital.

### 3.1 Population growth models

The simulation model needs to include a stochastic model representing the pandemic evolution to predict (simulate) the number of patients arriving at the health system. Because of their simplicity, we propose the use of PG models. A number of growth models are found in the literature, such as the models of Gompertz [59], Richards [60], Stannard [61], and logistic model and others [62]. They start with exponential growth but gradually decrease their specific growth rates. The equations that relate the number of cases in the population (infected, patients needing hospitalization, etc..) and the time are shown in Table 1.

**Table 1.** *Models considered for the data fitting and their equations*.

| Model | Equation |
| --- | --- |
| Logistic | $y = \frac{a}{[1 + \exp(b - cx)]}$ |
| Gompertz | $y = a \cdot \exp[-\exp(b - cx)]$ |
| Richards | $y = a\{1 + v \cdot \exp[k(\tau - x)]\}^{(-1/v)}$ |
| Stannard | $y = a \left\{ 1 + \exp \left[ -\frac{(l + kx)}{p} \right] \right\}^{(-p)}$ |

In this section, we carry out two statistical analysis for elucidating the adequacy of the PG models to represent and predict the evolution of the pandemic caused by the SARS-CoV-2 virus. The first analysis evaluates the capacity of the PG models to fit complete sets of real data representing positive cases registered in different countries. Specifically, PG models have been fitted to data coming from the 20 most-affected countries by COVID-19 on June 15, as it was recorded in Worldometer [63]. The parameter estimation of the PG models is done by minimizing the sum of squared errors. There are functions implemented in free software that perform this estimation of parameters, for instance, the *curve_fit()* function in the *optimize* module of SciPy in Python [64] or the *growthrates* package in R [65]. The fit quality is measured by the Mean Absolute Errors (MAE). Table 2 includes all MAE values calculated for each country and model. The best fits are marked in bold (differences less than 0,1% are not distinguished). Additional information in this table is the total population of each country and the total number of positive cases on June 15.

**Table 2.** *The 20 most-affected countries by COVID-19 until June 15. The last four columns show the MAE calculated for the fit with each of the applied models. Bold values represent the best scores*.

| # | Country | Population | Total positive cases (2020-06-15) | Logistic | Gompertz | Richards | Stannard |
| --- | --- | --- | --- | --- | --- | --- | --- |
| 1 | USA | 330,922,877 | 2,094,069 | 47,128.9 | **21,461.2** | 21,464.4 | 21,466.7 |
| 2 | Brazil | 212,496,348 | 867,624 | 3,182.3 | 3,245.1 | **2,754.7** | **2,754.7** |
| 3 | Russia | 145,932,063 | 528,964 | 5,667.4 | **1,688.7** | **1,689.0** | **1,689.2** |
| 4 | India | 1,379,418,901 | 332,424 | 1,392.9 | **537.9** | **538.0** | **538.2** |
| 5 | UK | 67,871,466 | 295,889 | 4,758.0 | **1,046.6** | **1,046.9** | **1,047.2** |
| 6 | Spain | 46,754,084 | 245,194 | 5,676.2 | **2,261.7** | **2,262.1** | **2,262.6** |
| 7 | Italy | 60,465,149 | 236,989 | 4,928.0 | **1,061.2** | **1,061.6** | **1,061.7** |
| 8 | Peru | 32,951,046 | 229,736 | 2,399.4 | **1,255.3** | 1,256.6 | 1,257.6 |
| 9 | Iran | 83,944,885 | 187,427 | 8,105.8 | **6,176.3** | **6,176.6** | **6,176.9** |
| 10 | Germany | 83,773,297 | 186,461 | 4,091.4 | **1,519.2** | **1,519.5** | **1,519.7** |
| 11 | Turkey | 84,299,464 | 178,239 | 5,335.7 | **2,418.3** | **2,418.8** | **2,419.1** |
| 12 | Chile | 19,109,226 | 174,293 | 1,132.5 | 1,601.2 | **1,068.5** | **1,068.5** |
| 13 | France | 65,267,844 | 157,220 | 3,396.4 | **1,547.0** | **1,547.2** | **1,547.3** |
| 14 | Mexico | 128,873,820 | 153,507 | **1,418.9** | 1,641.0 | 1,535.5 | 1,535.5 |
| 15 | Pakistan | 220,685,460 | 144,478 | 1,631.5 | 1,324.5 | **1,301.0** | 1,321.6 |
| 16 | Saudi Arabia | 34,788,836 | 127,541 | 1,709.0 | **814.9** | **814.9** | **814.9** |
| 17 | Canada | 37,728,057 | 98,776 | 1,494.6 | **331.0** | **331.1** | **331.1** |
| 18 | Bangladesh | 164,618,467 | 87,520 | 655.6 | **315.5** | **315.6** | **315.7** |
| 19 | China | 1,439,323,776 | 84,335 | 1,166.6 | 1,133.0 | **1,097.6** | **1,097.7** |
| 20 | Qatar | 2,807,805 | 79,602 | 417.7 | 369.2 | **263.6** | **263.6** |

These results show that similar fitting quality of Gompertz, Richards, and Stannard models, while the Logistic PG model underperforms the other three.

The second statistical analysis is designed to test the predictive capacity in the short-medium term of the PG models. The accurate prediction of future cases of the disease is crucial for the accurate prediction of the resources needed. Three experiments have been carried out to determine which model has a better predictive capacity. For each country, the dates in which the data exceed 25%, 40%, and 65% of positive cases registered on June 15 are selected. The rationale behind this choice is the following: 25% represents an early stage of the pandemic but with enough information to fit the curves; however, when few data are available, they lie in the exponential growth phase and lead usually to overestimations of the maximum of the function. This problem is accentuated in countries where the pandemic has a strong impact at the beginning, such as in Spain or Italy. In Section 3.2 it is explained how we deal with this problem. In addition, the values 40% and 65% are chosen to observe how each model predicts as more information becomes available.

The prediction of the fitted curves for the next 5, 10, and 15 days is assessed by calculating the MAE. These time horizons are considered as sufficient for the hospital managers to adapt extra resources for new needs. As new positive case data is added every day, and predictions are refreshed also every day, the long-term predictive capacity of the model will not be analyzed. The results of these analyzes are collected in Appendix A. On the one hand, we present fits to the curves until the selected days, obtaining an MAE for each model and country. On the other hand, the tables of the MAEs made in the predictions are shown. To facilitate the comparison of results, MAEs are normalized by the total number of positive cases on the selected days. From these results, we can conclude that the Gompertz model outperforms in predictive capacity the other PG models.

Table 3 summarizes the relevant information from all the tables in Appendix A. It indicates the number of countries in which each model is the best in terms of predictive capacity (as before, differences smaller than 0.1% have been considered equal). It is observed that the Gompertz model is the one that more accurately predicts future values, specifically in all the situations analyzed except for one. For this reason, the Gompertz model is recommended to predict new cases of COVID-19.

**Table 3.** *The number of countries in which each model is equal or better than the others in terms of predicting new positive cases for the next 5, 10, and 15 days. Bold values represent the best scores. *These values are of 18 countries because 2 of them have no real data for that period*.

| Model | 25% |  |  | 40% |  |  | 65% |  |  |
| --- | --- | --- | --- | --- | --- | --- | --- | --- | --- |
|  | 5 days | 10 days | 15 days | 5 days | 10 days | 15 days | 5 days | 10 days | 15 days |
| Logistic | 4 | 5 | 5 | 4 | 4 | 3 | 2 | 1 | 2* |
| Gompertz | **11** | **12** | **13** | **13** | **14** | **14** | 14 | **15** | **13\*** |
| Richards | **11** | 9 | 7 | **13** | 9 | 10 | **15** | **15** | 11* |
| Stannard | **11** | 8 | 7 | **13** | 9 | 10 | **15** | **15** | 11* |

### 3.2 Patient arrival pattern simulation

The Gompertz growth model can be used to simulate both the number of positive cases and the entry of patients to the health system. The parameters of the original equation of this model (Table 1) have a more mathematical than biological interpretation (*a*, *b*, *c* …), like most equations that describe a sigmoidal growth curve. Therefore, before using it in our modeling, it is convenient to carry out a transformation to easily interpret the results obtained in the fit. Zwietering et al. [66] rewrite the Gompertz growth model as it is shown in equation (1).

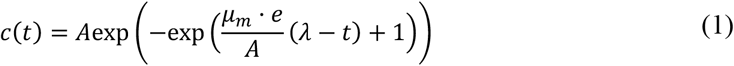

where,

- *e* = exp(1)
- *c*(*t*) is the cumulative number of positive cases or hospitalized patients until time *t*.
- *A* is a parameter of the growth model that represents the total number of positive cases or hospitalized patients at the end of the outbreak.
- *μ_m_* is the maximum specific growth rate of the curve.
- *λ* is the lag time, defined as the *t*-axis intercept of the tangent through the inflection point.

Once the curve *c*(*t*) is fitted, for example to cumulative hospitalized patients, it is used to simulate the number of new hospitalized patients of each of the following days. Let consider *t*_i_ and *t_i_*_+1_ two consecutive days in the future, the number of expected arrivals for the day *t_i_*_+1_ is calculated as *c*(*t_i_*_+1_) − *c*(*t_i_*). Then, the simulation model samples the number of hospitalized patients for that day *t_i_*_+1_ from a Poisson distribution with mean *c*(*t_i_*_+1_) − *c*(*t_i_*). This procedure is applied to simulate the hospitalized patients each future day. The Gompertz curve is fitted each time a new datum is observed.

At the first stages of the outbreak could not be enough data to fit the Gompertz model, overall in terms of hospitalized patients. In this case, we propose two alternatives that rely also on the use of PG models:

The first one consists of fitting the growth model to cumulative positive cases registered in a greater area, for example, the whole country when only a region of it is considered. Two factors are required to transform the predicted positive cases of the greater area into arrivals to hospitals in the subarea: the first one is a population factor that relates the population of both areas (e.g. the region of Navarre in Spain represents the 1.3% of the whole country population), and the second one transforms the new cases into patients requiring hospitalization (at first stages about 40% of detected cases needed hospitalization). Denoting by *f_g_* and *f_h_* both geographic and hospitalization factors, and by *C*(*t*) the fitted Gompertz curve to the positive cases in the bigger area, then *c*(*t*) = *f_g_f_h_ C*(*t*). Given that in this first period there is a data shortage, *f_h_* can be estimated by experts according to data published in the literature referred to the behavior of the disease in countries where it is more spread. This indirect technique could be applied also to the detected cases in the same area and apply only the *f_h_* factor.

Secondly, when the hospitalization cases begin to accumulate significantly (meaning the observed cumulative cases are starting its exponential growth) then we propose to fix (by expert opinion) the maximum population parameter (*A* in equation (1)) and to estimate the other two parameters by minimizing the sum of squared errors.

## 4 Modelling the patient flow

This section focusses on modelling the patient flow through the health system. In the first subsection, we describe all considered paths that a patient can take through the hospital. In the second subsection, we explain how the length stay of each patient is modelled.

### 4.1 Hospital patient path

COVID-19 patients can access the health system in different ways: a person can be diagnosed with COVID-19 illness in a primary healthcare facility, in the hospital emergency department, in a nursing home, after undergoing a SARS-CoV-2 test control (as a *Polymerase chain reaction* (*PCR*) test), etc. Depending on the severity of the illness the person is kept “isolated” at home or is admitted to the health care system as a COVID-19 patient, either in a hospital ward or in the ICU.

Treatment of COVID-19 patients requires dedicated resources, material, and personnel. We focus our study in the ward beds and ICU beds because they are scarce resources. Other necessary resources, as nurses and physicians, can be calculated from the number of required beds. The COVID-19 patient path through the hospital does not differ from other hospital patients. Fig. 1 shows the patient flow through the health system, highlighting the transitions between the hospital ward and the ICU. Patients can be admitted either into the ICU or into the hospital ward. A patient admitted to a hospital ward can be transferred to the ICU due to a health worsening. The discharge of the hospital ward is because of death or health improvement. Patients are discharged from the ICU to a hospital ward either because of death or because of health improvement.

**Fig. 1.**
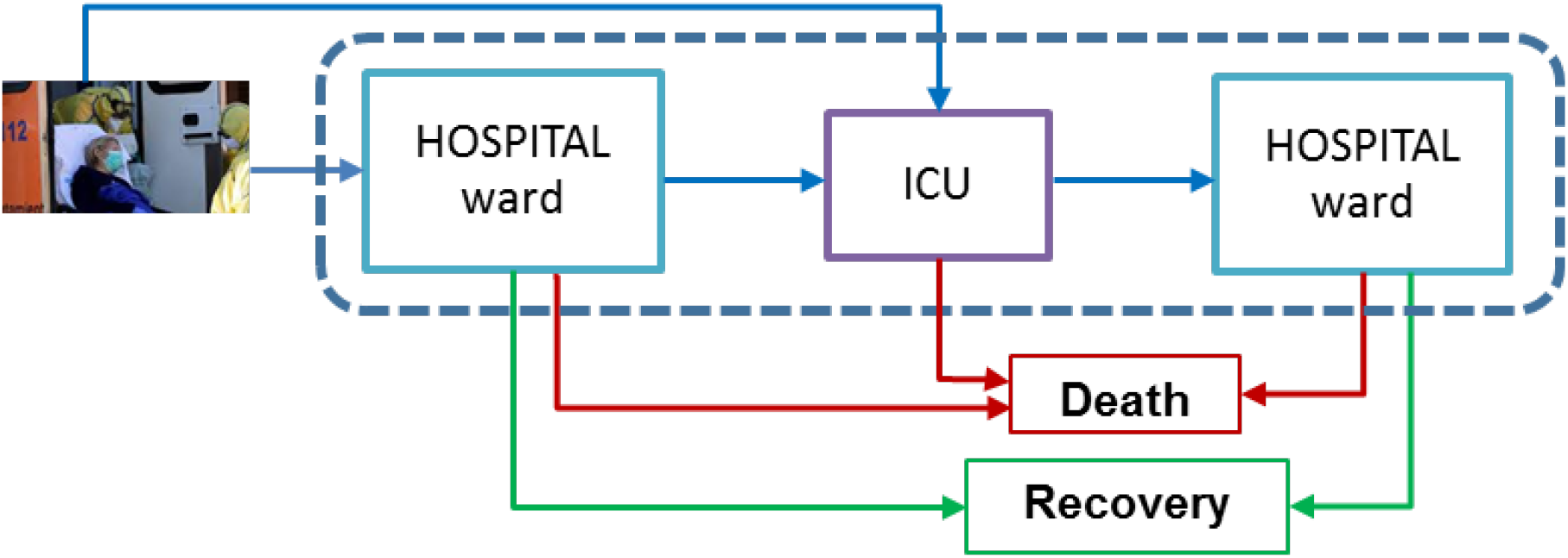
Representation of the COVID-19 patient flow in the health system.

### 4.2 Stochastic modelling of hospital length of stay

The following variables allow the modeling of the length of stay in the hospital:

- *X*_1_, the length of stay of a patient in the hospital ward not needing the ICU
- *Y*, the length of stay of a patient in the ICU.
- *W*, time spent by a patient in the hospital ward before being admitted to the ICU.
- *X*_2_, the length of stay of a patient in the hospital ward after being discharged from the ICU.
- *X*, the length of stay of a patient from its admission to a hospital ward or to the ICU until the discharge of the hospital facilities.

Besides, the following probabilities determine the patient-path through the healthcare facilities:

- *P*_1_, the probability that a patient is admitted to the ICU.
- *p_I_*_0_, the probability that a patient needing the ICU is admitted just after being detected positive. Then, 1 − *p_I_*_0_ is the probability that a patient is admitted to the ICU after being admitted first to a hospital ward.
- *p_WI_*, the probability that a patient admitted to a hospital ward ultimately needs to be transferred to the ICU. *p_WI_* = *p*_1_, (1 − p*_I_*_0_)/(1 − *p*_1_*p_I_*_0_)
- *p_IW_*, the probability that a patient is discharged from the ICU to a hospital ward. Then, 1 − *P_Iw_* is the probability that a patient dies in the ICU.

The estimation of both the probability distributions and the set of probabilities depends on the available data, which, in turn, depends on the pandemic development stage. During the first stages of the outbreak, when no patient hospitalization data or very few exist the model uses the triangular distribution family. The triangular is a popular family to estimate the time to accomplish a task because it embodies the idea of ‘three-point estimation’ where subjective judgment is used to estimate a minimum, a ‘best guess’, and a maximum value of the variable [67]. In the case of variable *X*_1_, these three values correspond to the minimum, most probable, and maximum times of hospitalization in a ward for patients not needing ICU, which are provided by expert opinion (hospital managers and the medical staff). Experts can rely on values reported in the literature corresponding to countries where the pandemic spread earlier (for example the China and the Italian situation are described in [68–71]).

As the pandemic progresses and there are more data on hospitalized COVID-19 patients, they can be used to estimate the probability distributions parameters. One main feature of these data is their high level of censorship; that is, only a small percentage of patients that have been admitted so far to the hospital ward and the ICU has been discharged after few weeks of the outbreak. This fact motivates the need for re-estimating the distribution fitting daily by adding the new data collected. The use of probability plots facilitates the selection of the parametric probability distribution family that best fits the data. The parameters of the selected family are estimated by fitting the updated health electronic recorded data on admission and discharge dates by the maximum likelihood method. Weibull and Lognormal distribution families have proved to be good probability models for the length of stay related variables, as we show in Section 7.

At the beginning of a new pandemic, there is insufficient knowledge of the illness and no effective treatments exist, as it has happened with the COVID-19 outbreak. As the medical and biological research progresses, new drugs and therapeutic protocols are discovered, which benefit the care of patients, affecting their length of stay in both hospital ward and ICUs. This observation reinforces the need for gathering every new data about patient admission and discharge to use them to update the estimation of the distribution parameters and bifurcation probabilities.

## 5 The discrete event simulation model

In this section, the mathematical modelling of the hospital dynamics is presented. First, we define a DES model to simulate the arrivals and the length of stay of patients. Then, a methodology is described that allows the simulation to start considering different levels of known information.

### 5.1 Entities, processes, events, and flowchart

DES model is defined by the set of state variables, which provide at any time a complete description of the simulated system, and the set of events, which modify the value of state variables. The simulation model represents the patient flow through the different ways of hospitalization, that is, the part inside the box with dashed lines in Fig. 1. In this subsection, we propose the state variables that describe the health system and a set of events grouped into three different categories.

Only one set of variables is considered in this model, which is composed of two variables, *H* = (*H_w_, H_I_*). These variables describe the number of patients hospitalized in hospital wards (*H_w_*) and the number of patients in the ICU (*H_I_*). Observe that the number of total patients admitted in the hospital *(N)* due to COVID-19 at time *t* is the sum of these two state variables (*N* = *H_W_* + *H_I_*).

There are three different types of events that modify through time the value of state variables. They have been classified according to how the value of *N* changes, if there is an increase, a decrease, or if its value does not change after the event occurs. The first set of events *E_A_* are associated with the patient’s arrival times classified in hospital arrivals and ICU arrivals. In this group, we only consider external arrivals, i.e., patients who are not in the hospital and directly arrive after being detected with the disease. These arrivals occur as we mentioned in Section 3.2 distinguishing with a percentage what type of patients they are.

The second category of events *E_B_* produces a decrease in the state variables. They are related to the end of the patient hospitalization time. In wards, we consider both discharges for improvement and death, but in ICU only death cases are included here. In this way, these events only reduce the values of *H_W_* or *H_I_*, and therefore the value of *N* decreases. In Section 4.2 it is explained how the length of stay is assigned to each patient, and therefore, the discharge times.

The third set of events *E_c_* is associated with the transfer of patients between the hospital wards and the ICU, which can be bidirectional (see Section 4.2). On the one hand, a patient who is in the hospital may worsen his health of status and have to be transferred to the ICU. On the other hand, ICU patients who improve and are discharged do not go directly home but are first treated in wards. Observe that in both cases, only the values of *H_W_* and *H_I_* are affected, but the value of *N* remains constant since with these actions neither patients enter nor leave the system. Fig. 2 outlines the simulation model of the health system.

**Fig. 2.**
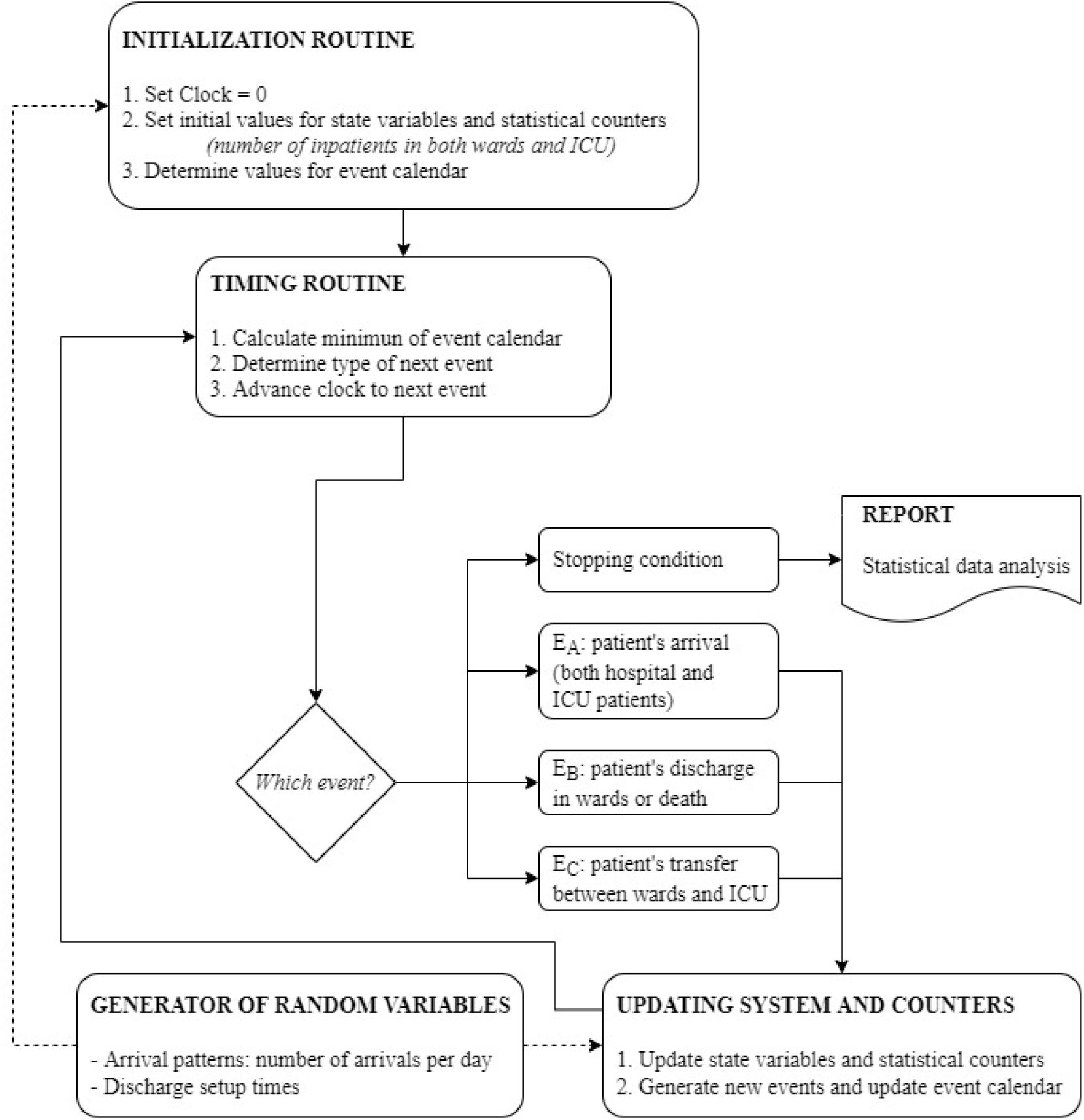
Flow diagram of the health system simulation model. Three types of events are considered (patient’s arrival, patient’s discharge, and patient’s transfer).

### 5.2 How to get the simulation started?

The purpose of the simulation model is the short-term necessary resources prediction. Therefore, the precision of the predictions strongly depends on the model accuracy in both representing the health system’s initial state and the initial time evolution of the already occupied resources. Usually, this last aspect of the mathematical modeling is not such important when the simulation is intended to investigate the behavior of systems in the long term, in its stationary state, which is usually independent of the system’s initial state. However, when the simulation is used as a prediction tool to support tactical decisions, the initial state of the simulation model and the system dynamic at the initial moments are the main factors to determine the health system state in the near future. The number of patients in the hospital wards and in the ICU, as well as the available information about the time since their admission, define the initial state of the health-system simulation model.

The simulation clock is set to zero at the time of the EHR-file’s last update. Before that time, it is the past that can be reproduced by the simulator by reading the records in the EHR file, and ahead, it is the future that needs to be simulated. The transition from the past to the future is done at the moment the simulation starts, which requires to initialize the event calendar [67]. There are three types of events, admission to hospital, transferring between hospital facilities (ward and ICU) and discharge from hospital. The simulation of the event type “arrival of a new patient” was explained in Section 3.2. Now we expose how to simulate the other two types of events to those patients already admitted in the hospital at time zero.

The simulation of these events depends on the available information about the patients. Basically, the main difference is whether there is access to patient-level information or only aggregated information about daily admissions and discharges is available. Zero time for the simulation model is considered to be the end of the *kth* day of the pandemic. That is, the simulation model begins to simulate what will happen from day *(k* + 1)*th*, using the information collected during the first *k* days of the pandemic and considering patients admitted to the health system at the end of the *kth* day as the initial state. The following notation, together with the notation introduced in Section 4.2, is used in the subsequent analysis:

*H_Wk_*: the number of hospitalized patients in hospital wards at the end of the day *k*.
*H_Ik_*: the number of admitted patients at the ICU at the end of the day *k*.
*I_j_* and *O_j_* number of new patients admitted, and number of patients discharged the day *j*, respectively.

They will refer to the hospital wards or to the ICU depending on the context of the analysis. *u*: an observation from a Uniform [0,1] random variable.

A. **Information at the patient level available: The admission date of each hospitalized patient is known**. Therefore, for each currently hospitalized patient the time *s_t_* already spent at the hospital ward or the ICU is known. ***Patient admitted to the ICU:*** The discharge date is calculated by sampling from the random variable *Y* conditioned to a stay longer than, the number of days already spent at the hospital. Let *y_t_* be a value sampled from the conditional distribution *Y/Y* > *s_i_*, then the value *y_i_* − *s_i_* is the ICU discharge time simulated for patient *i*. ***Patient admitted to a hospital ward:*** A patient occupying a ward bed can ultimately be discharged from the hospital or be transferred to the ICU. The probability that a patient hospitalized for days is transferred in the future to the ICU, denoted by 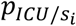, is calculated by using Bayes theorem:

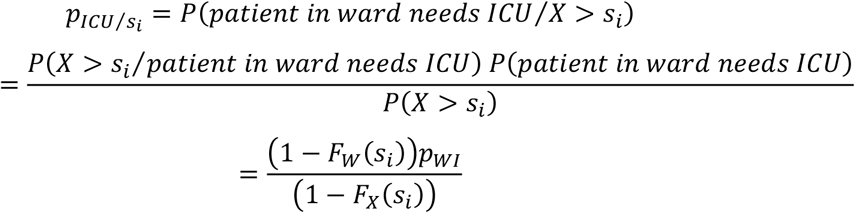 Therefore, for each patient *i*, a random number *u* is used to classify each patient into one out of two categories: patient to be transferred to ICU, when 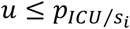, or patient to be discharged from the hospital, when 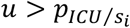. In the case of a patient transfer to the ICU, the event transfer time is simulated by sampling a value *Wi* from the conditional distribution *W/W* > *s_i_*. The value *w_i_* − *s_i_* is the simulated transfer time from hospital ward to the ICU. Similarly, the discharge time from the hospital ward is simulated by sampling a value *x_i_* from the conditional distribution *X*_1_/*X*_1_ > *s_i_*, then the value *x_i_* − *s_i_* is the simulated discharge time.
B. **The series of new admitted patients *I_j_, j* = 1,…, *k* and the number *H_Wk_* of hospitalized patients are known at the moment of running the simulation model**. However, individual arrival dates are not known for the admitted patients and the method developed for the case A can not be applied. Let *P_ij_* be the probability that a patient admitted the day *i* is discharged the day *j*, and *q_ij_* the probability that a patient admitted the day *i* is discharged later than day *j*. The end of day *k* is the present time at which the simulation model is run. The probability that a patient arriving the day *i* is still hospitalized is *q_ik_*. Every past day *i, i* = 1,*k, I_i_* new patients are admitted, which account for 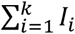 patients of which *H_Wk_* are still hospitalized. The method sketched in Fig. 3 selects *H_wk_* patients out of 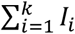 by a sampling process without replacement that uses the set of probabilities {*q_ik_*; *i* = 1,…, *k*}. The application of this method results in the simulation of the admission date of each admitted patient. Therefore, the simulation of the discharge and transfer events can be performed according to the method of case A. The probabilities *q_ik_* are calculated for patients admitted in the ICU from the probability distribution of variable *Y*, and for patients admitted to a hospital ward from the probability distribution of variable *X*.
C. **For each day *j, j* = 1,…, *k*, the number *I_j_* of new admitted patients and the number *O_j_* of discharged patients are known**. We use this information (see Fig. 4) to estimate for each patient the probability of being one of the *H_Wk_* admitted patients at the end of day *k* and the day of admission *i*. Once these probabilities are calculated, the procedures exposed in cases A and B can be applied. Using, as before, the notation *p_ij_* for the probability of a patient being admitted the day *i* and discharged the day *j*, and *q_ij_* for the probability of a patient being admitted day *i* and discharged later than day *j*, then the expected number of discharge patients the day *j* is 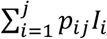 and the observed number *O_j_*. Let *g_j_* be the factor verifying *g_j_* 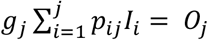, so that *n_ij_ = g_j_p_ij_I_i_* is the updated expected number of patients arriving the day *i* that are discharged the day *j*. Similar calculation is performed for the number of admitted patients at the end of day *k*: 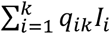 is the expected number of admitted patients at the end of day *k* that arrived the day *i* and *H_Wk_* the actual number of admitted patients. Let 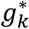 be the factor verifying 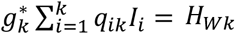, and therefore 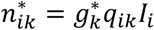 is the updated expected number of patients arriving the day *i* that still are hospitalized at the end of day *k*. The estimated new expected values *n_ij_*, *j* = *i*,…, *k and* 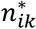 are used to rescale the probabilities *q_ik_*: let *y_t_* the factor satisfying that 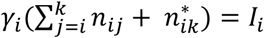, that is, 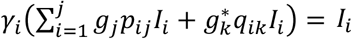 and then, 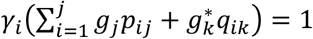,. Therefore, 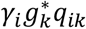 is the updated estimation of the probability that a patient arriving the day *i* is still hospitalized at the end of day *k*.
D. **Only the number *H_Wk_* of hospitalized patients is known**. This case happens when the forecasting of daily hospitalized patients is performed from the series of aggregated positive cases in the local area, *c(t)*, and a hospitalization factor *f_h_* is applied. In this situation the arrival date of the patients currently admitted at the hospital (end of day *k*) is simulated by first simulating the arrival of patients each day before (*i* = 1,…, *k*) and then by applying the method of paragraph B. The number of hospitalized patients each day *i* = 1,…, *k* is simulated by sampling values from a binomial distribution with parameters *c(i)* − *c*(*i* − 1), the total number of new positive cases in day *i*, and *f_h_*, the probability that a positive case needs to be hospitalized.

**Fig. 3.**
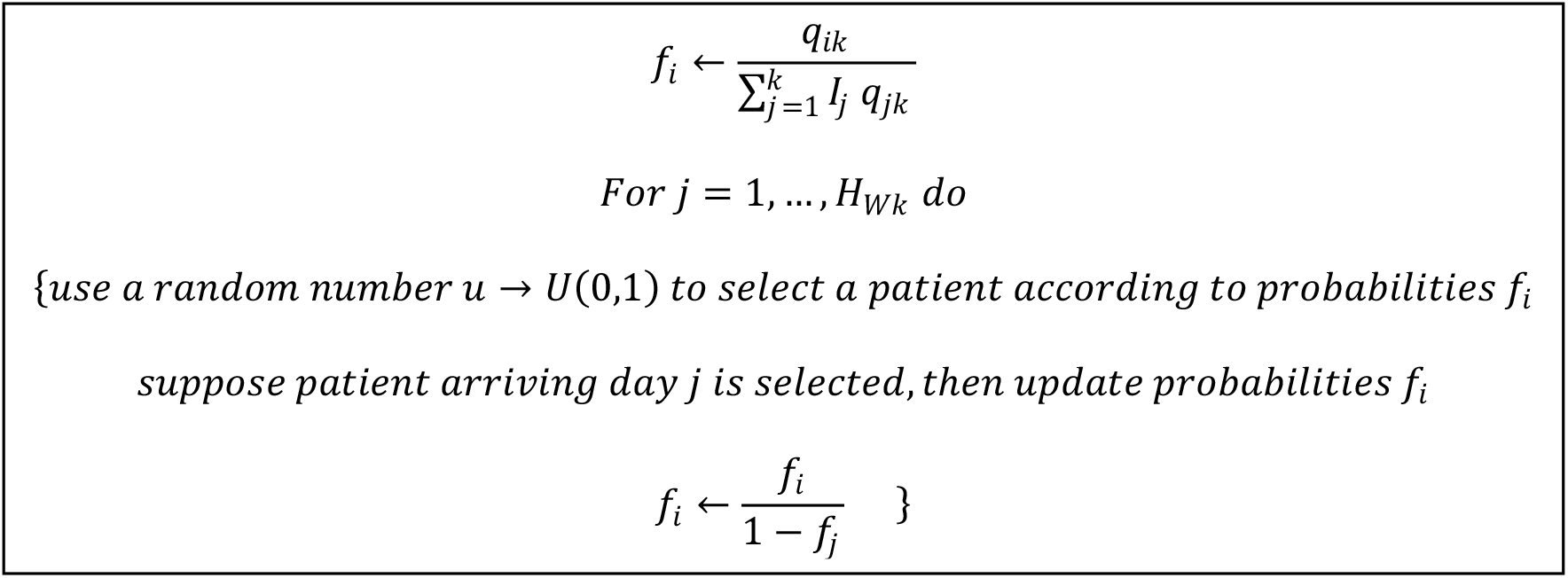
The algorithm implemented to select which patients are hospitalized at the beginning of the simulation. For the ICU the algorithm is the same but referring all the parameters to the ICU (arrivals, bed occupation…).

**Fig. 4.**
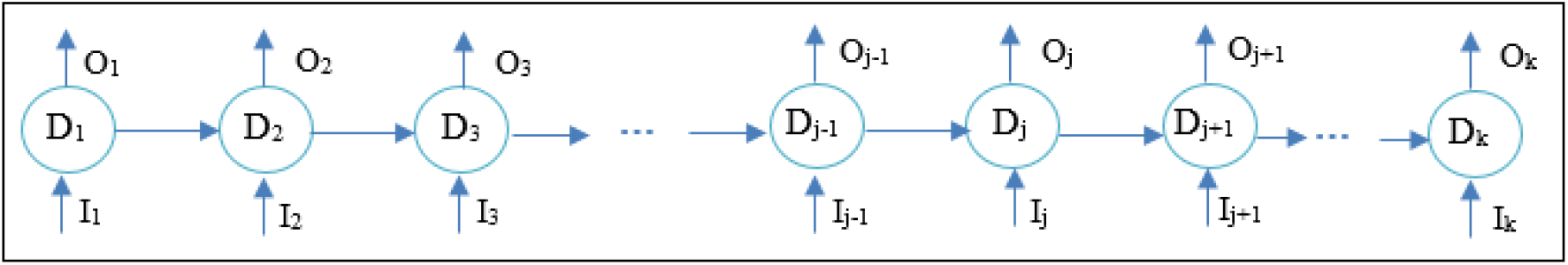
Diagram that illustrates the number of new admitted patients (I_j_) and the number of discharged patients (O_j_) per day (D_j_).

Once a discharge time or a transfer time between ward and ICU has been simulated for each hospitalized patient, the event calendar records those times, together with the arrival time of the next COVID-19 patient. The fitted Gompertz curve provides the number of patients arriving each day. These arrivals can be uniformly distributed during the next 24 hours or following a non-stationary pattern when, for example, a significant decrease of arrivals occurs at night. The clock of the simulation is advanced from zero to the minimum of the times recorded in the event calendar vector.

## 6 The simulator

The simulator developed for this work is presented in this section. The types of inputs required to start the simulator are described first, depending on the level of information known. Subsequently, the results obtained with this tool are shown graphically.

### 6.1 Input data

The developed simulator has a lot of flexibility to simulate a large number of scenarios and in different conditions. This allows medical staff to adapt the model to their particular hospital while testing different results by modifying some parameters. There are two types of input data to the simulator, depending on whether they are input parameters that are configured on the simulator screens before predictions are made or if they are external files to the program that the simulator needs to read to perform the simulation.

#### Simulator parameters

The simulator allows the user to modify the factors mentioned in Section 3.2 (*f_g_* and *f_h_*). As explained in this section, in the early stages of the pandemic, it can be difficult to fit the curve of hospitalized patients directly due to the scarcity of data. In that phase, the cases of a larger region are fitted, and the predictions are scaled with both factors. These factors can be introduced following the experts’ criteria (overall *f_h_*) or based on the proportion of the population when scaling cases of positives from a larger region to a smaller one. When fitting the hospitalized patient curve, the factors mentioned must be 1.

Another parameter to introduce is the probability that a patient who arrives at the hospital needs special care in the ICU (*p_ICU_*). The percentage of such patients who are directly admitted is also indicated (*p_ICU_*__0_), distinguishing them from those who spent a few days in wards before (*p_ICU_*__1_). In turn, it is possible to define the distribution that the length of stay of the COVID-19 patients follows (Weibull, lognormal …). For each selected distribution, the simulator allows the user to configure its parameters.

Finally, the simulator allows the user to select one of the four types of simulation outlined in Section 5.2 (A, B, C, D), depending on the amount of information available to the user. He or she should consider the simulation type selected to provide the correct input files.

#### External files

One of the input files required by the simulator is the accumulated historical series, either of positive cases or patients admitted to the hospital. One series or another will be used depending on the phase of the pandemic or the amount of information available. These series are used to obtain the best fitting Gompertz curve, which allows the patient arrivals pattern to be estimated.

The other input files are related to the type of simulation to be run. If full information is required (A), the simulator is fed by an input data file obtained from the hospital electronic health record (EHR) system. The information needed for each COVID-19 patient arriving at the health system is the dates of hospital admission and discharge and the dates of ICU admission and discharge. If a field is empty, then the associated event has not occurred. For example, an already discharged patient from hospital with empty fields in the ICU fields means that this patient did not need intensive healthcare. A patient will be admitted in the health system at the beginning of the simulation when they have an admission date, but their discharge date field is empty.

The input data file records the information of all COVID-19 patients from the first day of the outbreak to the present day. Therefore, in this modality, the simulator can reproduce the occupancy of the hospital and ICU from the beginning of the outbreak until the present, and at this moment it is known the number of patients in the health system (in all hospitalization modalities) and the date of admission of those patients. This information is used to start the simulation run as it is explained in Section 5.2.

Finally, the simulation could also be run without specifying all the disaggregated information of each patient in the health system, just by knowing the number of patients admitted in wards and in the ICU (D). Or knowing, in addition to this, both the number of daily admissions and patient discharges (C) or only the admissions (B). These series are provided as external input files. In these cases, the simulator is not able to reproduce the past and get a very accurate representation of the health system at time zero of the simulation clock, but it can be estimated.

### 6.2 Simulation output

During the simulation, the program generates arrivals and discharges of patients of both the hospital and the ICU. The system evolves and the number of necessary beds is recorded at any time. In this simulation model, there are two sources of randomness. On the one hand, the number of patients arriving at the system (hospital and ICU), and on the other, the length of stay of patients, according to the distributions mentioned in Section 4.1. In each program run, the configured scenario is simulated many times (thousands), and thus a distribution of the number of beds needed each day is obtained since, in each simulation, the occupancy curve varies. The simulator calculates percentiles, which are stored in an Excel file, specifically the 5^th^ percentile (P5), the 50^th^ percentile (P50), and the 95^th^ percentile (P95). Also, these percentiles, which represent confidence bands, are plotted on a graph.

Fig. 5 shows the graphic outputs that are obtained with the program. On the left side, the necessary hospitalization beds are observed, while on the right side the same graph is shown but for the ICU occupation. The green-colored line represents the real evolution of the occupation, and the black dot indicates the Simulation Starting Point (SSP), that is, the moment from which it is simulated.

**Fig. 5.**
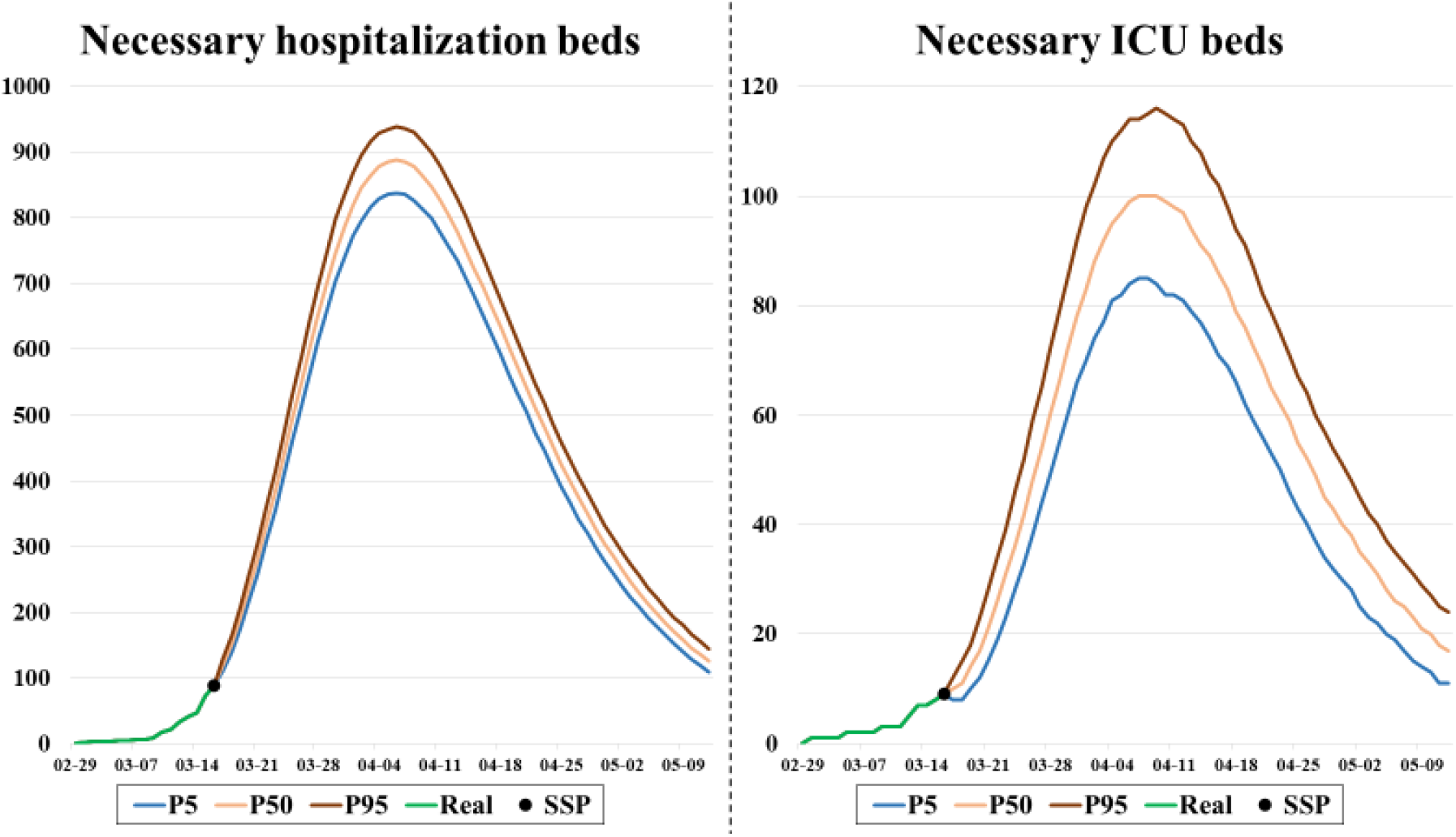
Number of necessary hospitalization and ICU beds respectively for the following days. From the SSP, 3 lines corresponding to the 5^th^, 50^th^, and 95^th^ percentiles are plotted.

## 7 Application to real cases

In this section, a case study is presented, in which the explained methodology has been applied to a region of Spain (Navarre). Firstly, we briefly introduce how the virus has affected Navarra globally, and then, some predictions of beds obtained with the program in different moments are presented and compared with the real bed occupancy curve.

### 7.1 Incidence of COVID-19 disease in Navarre (Spain)

Navarre is a small community of the north of Spain. Its population is around 650,000 inhabitants, with more than a half gathered around the capital (Pamplona) and its surroundings, an area that represents 5% of the total surface. With this distribution of population, the Health Services of Navarre has a main hospital located in Pamplona, with more than 1,000 available hospital beds, and two secondary hospitals in two of the most populated cities (Estella and Tudela). In total, the public health system in Navarra has 1,466 hospital beds and 45 ICU beds, with the possibility of increasing these quantities if necessary.

Navarre was among the five Spanish autonomous communities with a higher cumulative incidence rate of COVID-19 confirmed cases at the beginning of the pandemic, according to the data collected by the Government of Navarre [72]. By May 12, 7,752 COVID-19 cases (11.9 per 1,000 inhabitants) had been confirmed in Navarre, and among them, 1,704 had been admitted to public hospitals (2.6 per 1,000). Fig. 6 shows four graphs with daily and cumulative data about positive cases and patients who have been admitted to public hospitals in Navarre. It is observed that time trends peaked on March 25 and 27 and decreased after April 1 and 2.

**Fig. 6.**
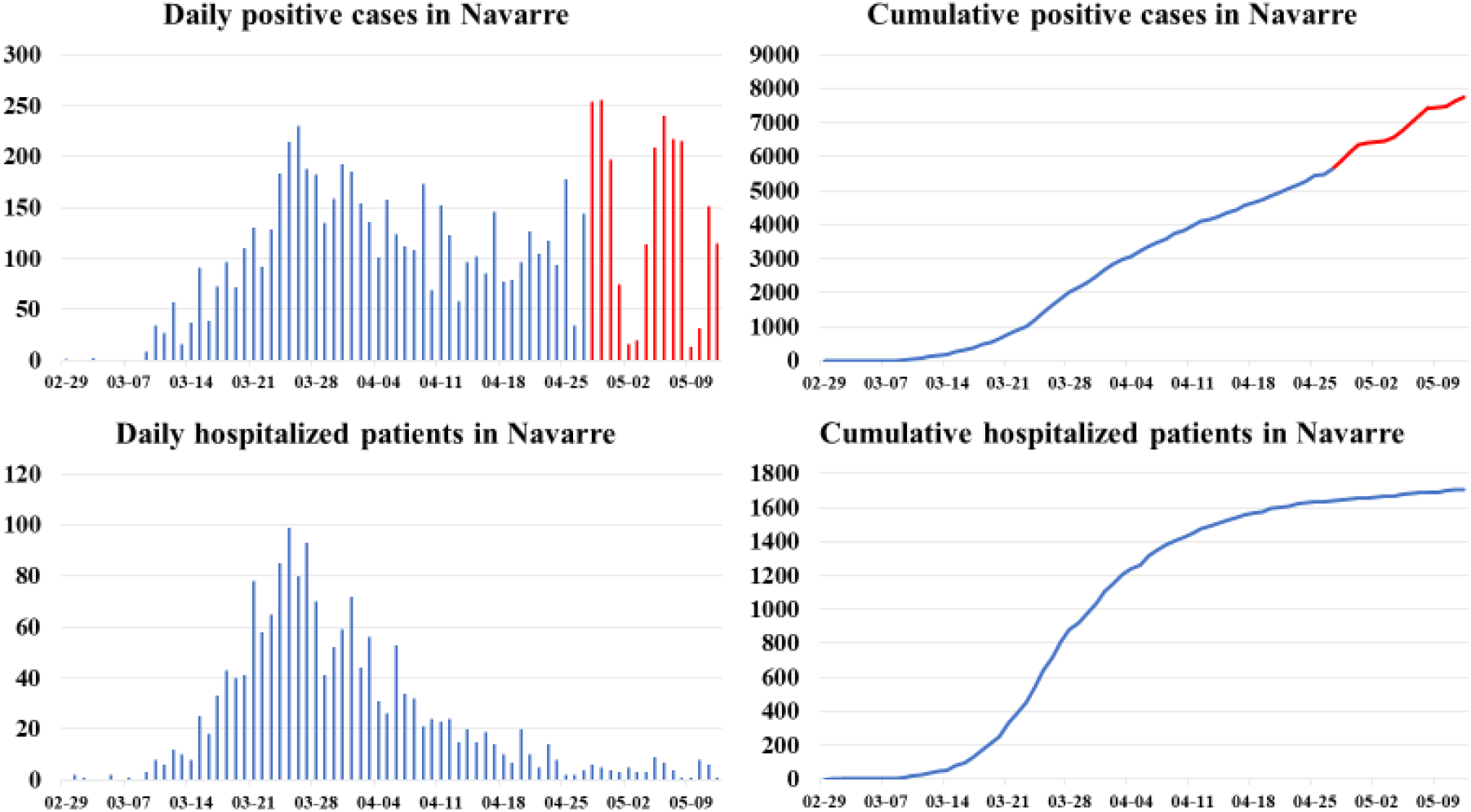
Daily and cumulative data ofpositive cases and hospitalized patients recorded in Navarre from February 29 to May 12.

Observe that as of April 28 there seems to be a rebound in the number of positive cases, (see red-colored values), which are not reflected in the number of hospitalized patients. This is because in the first stage of the pandemic the system’s ability to perform tests was limited, and symptomatic patients were those who approached the system. However, as cases have decreased, the health system has reached out to patients to prevent further outbreaks, and far more cases of asymptomatic individuals have been detected than previously accounted for. This shows that when adjusting the Gompertz model based on positive cases, there are parameters, such as the hospitalization factor *(f_h_*), that vary dynamically and must be updated.

### 7.2 Predictions of both hospitalization and ICU bed occupancy

#### 7.2.1 Fitting the cumulative positive cases

In the early stages of a pandemic, there is a scarcity of data on both the number of people infected with the virus and hospitalized patients, as well as in the length of stays observed in the hospital. Therefore, the involvement of experts is very important and necessary to estimate these parameters at these stages. In this way, a first approach can be made to simulate the evolution of the pandemic in the following days.

Navarre was one of the first communities in Spain to be infected, so at the beginning, the historical series of people infected with the virus was used to estimate the patient arrival pattern. Another possibility would be to use the cumulative positive cases of Spain and apply the appropriate geographic factor *(f_g_*). The problem that usually appears at the beginning of the outbreak is that the trend may lead to an exponential growth of the series. To avoid this, the total number of expected positive cases (*A* in equation (1)) is fixed, to obtain a realistic output from the Gompertz growth model fit.

Fig. 7 shows different results after applying the Gompertz growth model fit to the cumulative positive cases in Navarre. In the early stages of the pandemic (03-16), two curves can be seen, one of them obtained using the cumulative positive cases of Spain (03-16(Spain)). In addition, three more fits made on March 31 (03-31), April 15 (04-15), and April 30 (04-30) are exposed. Note that the last values in the series are not predicted very accurately and this is due to what was discussed in Section 7.1. These results demonstrate that accounting for positive cases has changed.

**Fig. 7.**
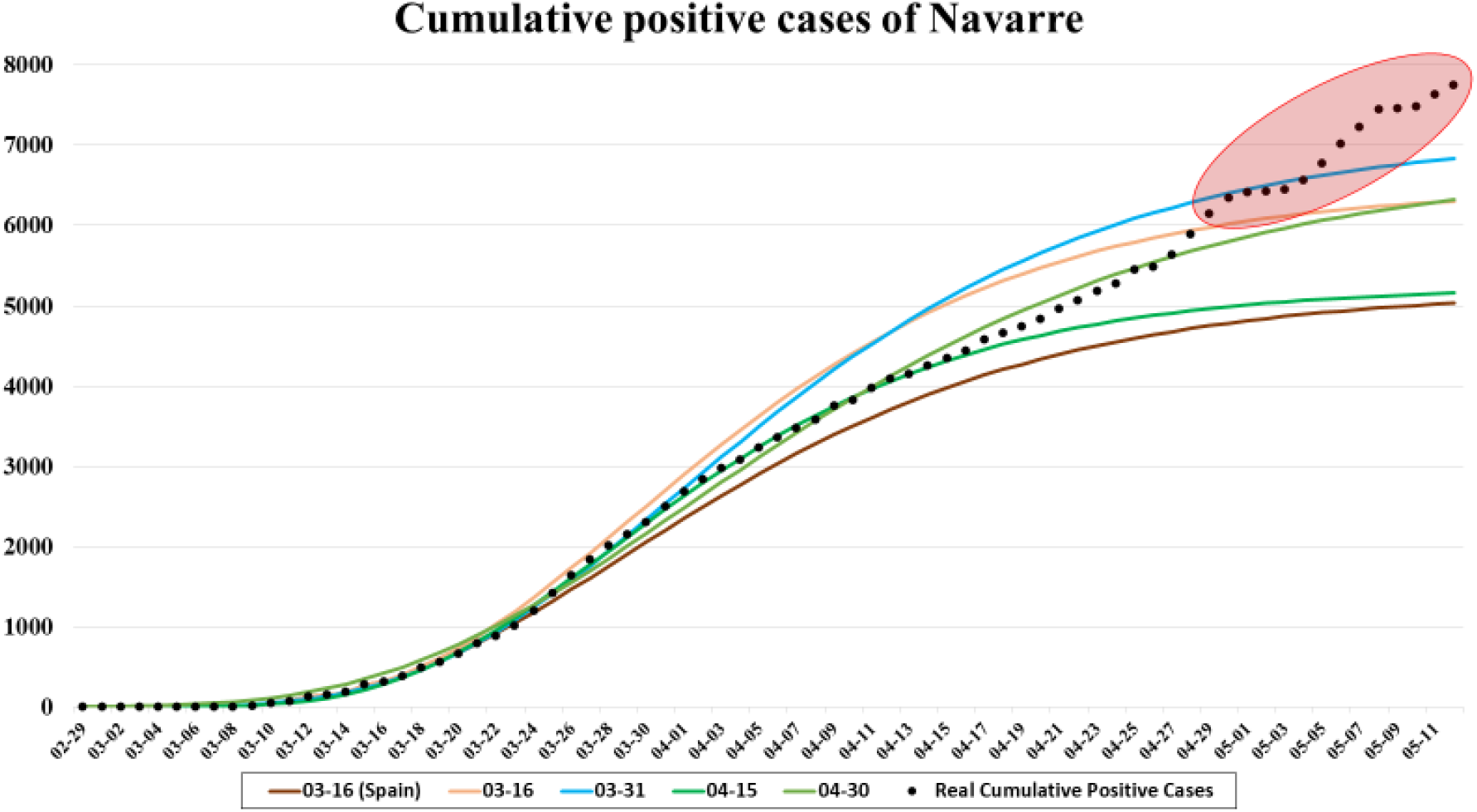
Cumulative positive cases of Navarre from February 29 to May 12 and different fitted curves obtained from the Gompertz growth model.

The first two curves on March 16 were obtained after setting parameter *A*. For the case of Spain, *A* = 175,000 was established according to the cumulative incidence observed in China and reported in the bibliography, and comparing the evolution with Italy, where the pandemic outbroke a few weeks earlier. Then *f_g_* scales the curve. The criterion for choosing its value was the rate of positive cases between Navarre and Spain at that time (3%) rather than the rate of populations between both regions (1.3%). In the case of the cumulative positive cases in Navarra, *A* = 6,500 was set (1% of the total population of Navarre), since the population in this region is more concentrated than in the rest of Spain, and this makes the infection more aggressive.

At the initial stage of the outbreak when there is not enough data to estimate the length of stay of patients, a triangular distribution is used. Experts fix the minimum, maximum, and most probable time as 10, 18, and 13 days respectively. In addition, a hospitalization factor (*f*_g_) must be applied (0.35 for hospitalized patients and 0.04 for ICU patients), because the arrival pattern is based on positive cases. Fig. 8 shows both the prediction of the number of beds occupied in the hospitals and in the ICUs of Navarre simulated on March 16 for the next days. On the upper side, the predictions based on the cumulative positive cases of Navarre are observed, while those on the lower side are based on the cumulative positive cases of Spain. In each graph, the real evolution of bed occupancy in each area is shown in green.

**Fig. 8.**
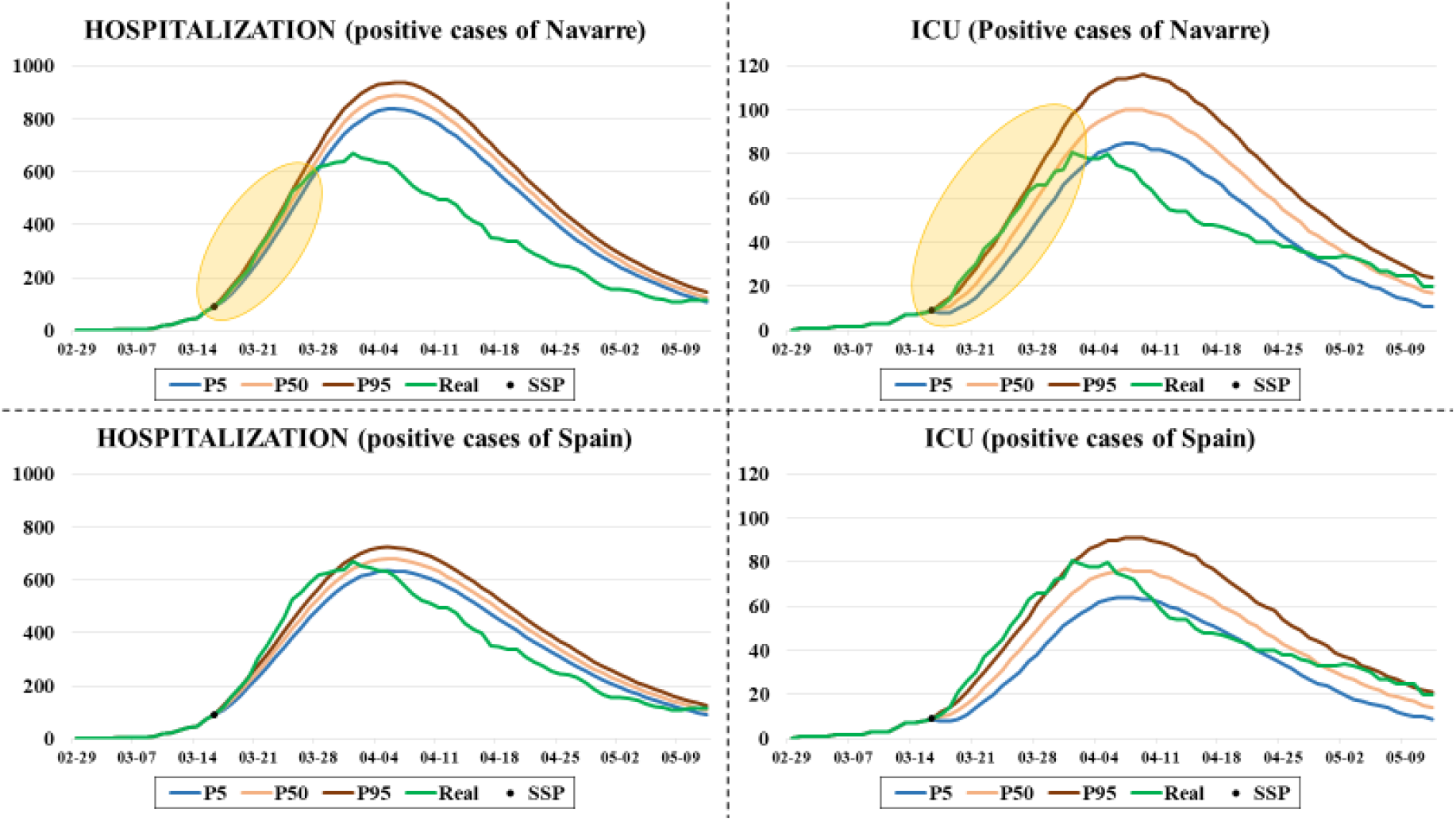
Comparison between the predictions made on March 16 for the number of beds occupied in the hospitals and the ICUs of Navarre and the real occupancies. The cumulative positive cases of Navarre have been used for predictions in the two graphs at the top, while the cumulative positive cases of Spain in those at the low.

Note that the most important predictions for the medical staff are for the short-medium term (yellow-colored area in Fig. 8), and the results obtained are very similar to the real ones concerning both hospital and ICU occupancies. Fig. 9 shows a zoom of these parts of the graphs. The simulator’s ability to obtain accurate predictions for the next 15 days, even in the early stages of the pandemic, is demonstrated here.

**Fig. 9.**
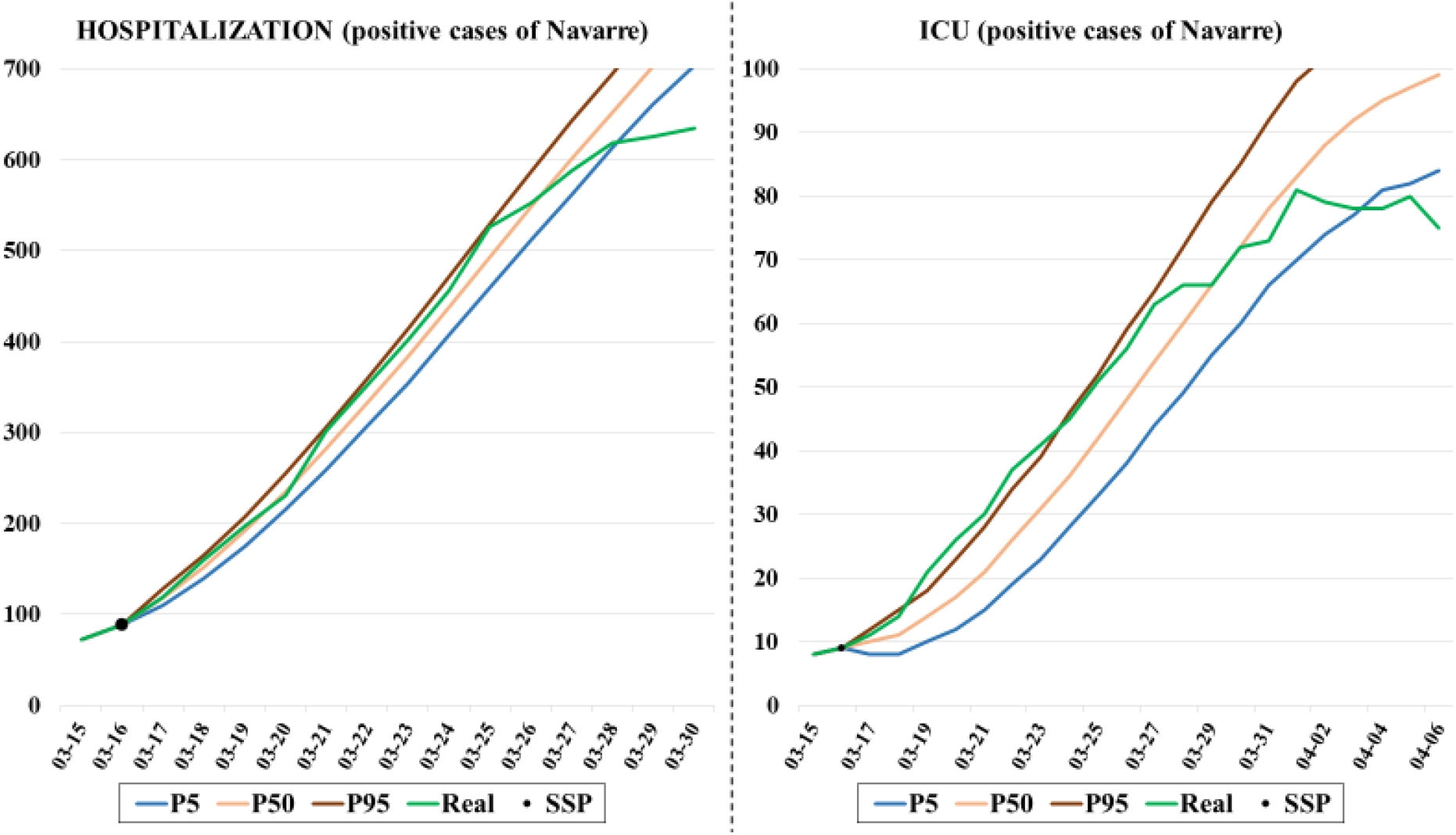
Expanded part of the predictions (for both hospitalization and ICU) made on March 16 for the next 15 days using the cumulative positive cases of Navarre.

#### 7.2.2 Fitting the cumulative hospitalized patients comparing levels of information

As the pandemic spreads, the amount of information increases, which makes it possible to improve the simulation model. Since March 26, the arrival pattern is calculated from the series of patients admitted to the hospital. In the first few days, it was required to fix the total number of expected hospitalized patients (*A*), as with the positive cases, due to the curve fitted by the Gompertz growth model increases exponentially. According to experts, about 2,000 patients were expected to be admitted to public hospitals during the pandemic. Fig. 10 shows different results after applying the Gompertz growth model fit to the cumulative hospitalized patients in Navarre. It can be seen that the curve obtained on March 26 overestimate hospitalizations, so it was replaced in the simulation by that in which the maximum is set at 2,000 (03-26 (2,000)).

**Fig. 10.**
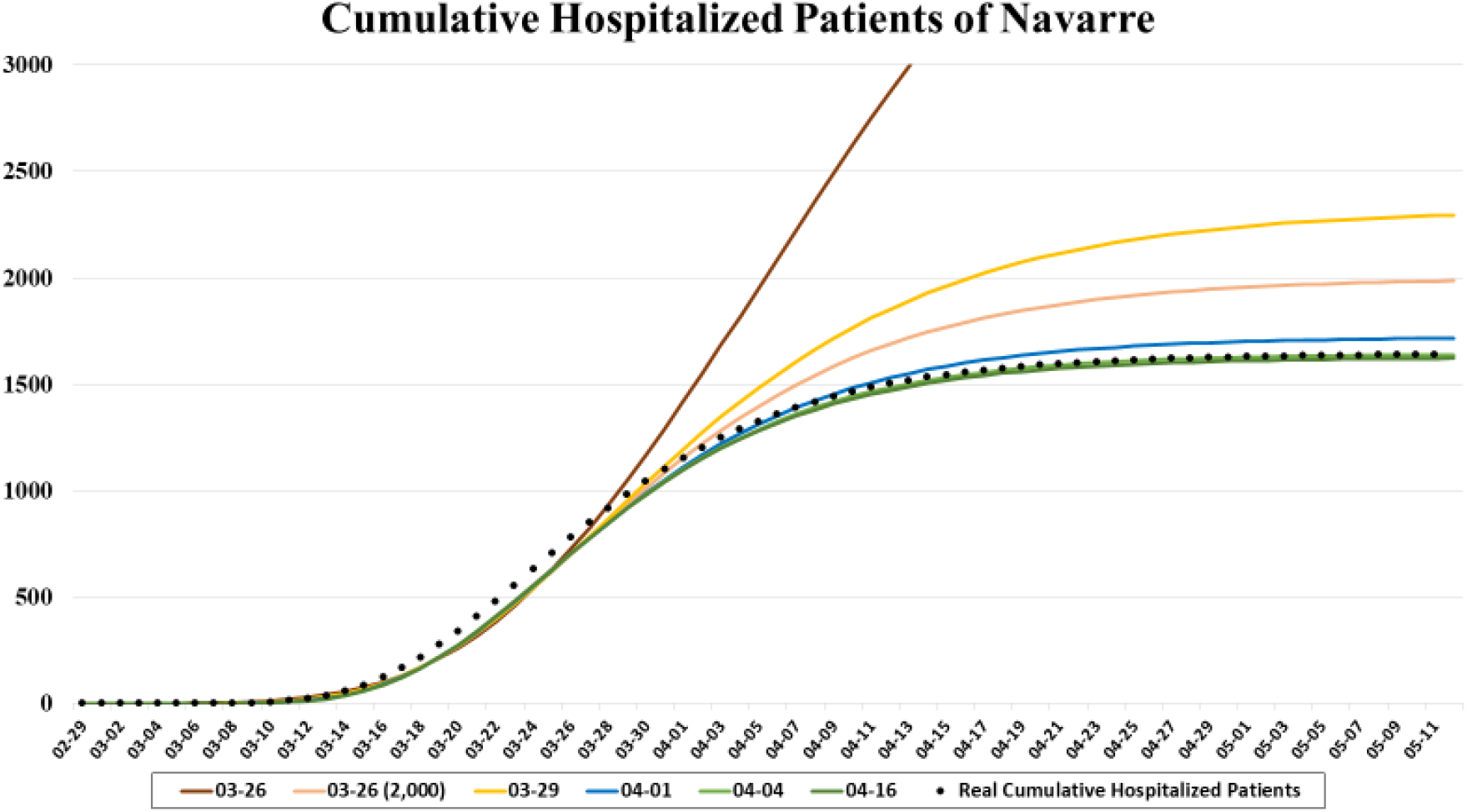
Cumulative hospitalized patients of Navarre from February 29 to May 12 and different fitted curves obtained from the Gompertz growth model.

Regarding the length of stay of patients both in hospital and ICU, there were already enough data to estimate them every day, as it is explained in Section 4.2. At the beginning of the pandemic, the length of stay of patients showed a reasonable fit to a Weibull distribution, although, as time goes by and more data has been obtained, the lognormal distribution has shown to be a better model. Each time the data was analyzed, the parameters of the distribution were configured for the simulation. Table 4 lists those parameters used in the simulations at different moments of the pandemic, as well as some of the variables entered manually in the simulator defined in Section 4.2.

**Table 4.** *The parameters used in the simulation at different moments during the pandemic. The distributions of the different lengths of stay considered as well as the probabilities related to ICU patients are shown*.

| Date | $X$ (days) | $\bar{X}$ (days) | $Y$ (days) | $\bar{Y}$ (days) | $Z$ (days) | $\bar{Z}$ (days) | $p_I$ | $p_{I0}$ | $p_{IW}$ |
| --- | --- | --- | --- | --- | --- | --- | --- | --- | --- |
| 03-26 | Weibull<br>(1.223,<br>18.470) | 17.291 | Weibull<br>(1.450;<br>22.707) | 20.589 | Weibull<br>(2.337,<br>3.802) | 3.369 | 0.075 | 0.390 | 0.444 |
| 03-29 | Weibull<br>(1.204,<br>18.489) | 17.376 | Weibull<br>(1.450;<br>24.663) | 22.363 | Weibull<br>(2.424,<br>4.016) | 3.560 | 0.046 | 0.310 | 0.308 |
| 04-01 | Weibull<br>(1.261,<br>15.421) | 14.334 | Weibull<br>(1.450;<br>25.423) | 25.423 | Weibull<br>(1.670;<br>4.410) | 3.939 | 0.043 | 0.310 | 0.353 |
| 04-04 | Weibull<br>(1.030,<br>15.000) | 14.821 | Weibull<br>(1.430;<br>28.677) | 26.053 | Weibull<br>(1.691;<br>4.445) | 3.967 | 0.042 | 0.272 | 0.36 |
| 04-10 | Weibull<br>(1.050;<br>14.914) | 14.627 | Weibull<br>(1.500;<br>23.948) | 21.619 | Weibull<br>(1.652;<br>4.436) | 3.966 | 0.042 | 0.272 | 0.500 |
| 04-16 | Weibull<br>(1.000;<br>14.844) | 14.844 | Weibull<br>(1.400;<br>24.304) | 22.151 | Weibull<br>(1.646;<br>4.385) | 3.921 | 0.040 | 0.309 | 0.619 |
| 04-26 | Lognormal<br>(2.275;<br>1.070) | 17.241 | Weibull<br>(1.050;<br>26.006) | 25.507 | Weibull<br>(1.464;<br>4.565) | 4.134 | 0.042 | 0.331 | 0.628 |

With all this information updated day by day, and the curves shown in Fig. 10, it is possible to simulate the evolution of the bed occupancy from a specific day. Fig. 11 shows two types of simulations carried out on March 26 and April 4. The difference between them lies in the amount of information used for the simulation. On the one hand, all the information was available, such as series of hospitalized patients and detailed information of the patients admitted (dates of admission and discharge of each patient, admission to the ICU, etc.). On the other hand, only the information about the series of hospital and ICU admissions, and the number of patients admitted at the beginning of the simulation (*H_Wk_* y *H_Ik_*) were used. These assumptions correspond to scenarios A and B respectively, which are developed in Section 5.2.

**Fig. 11.**
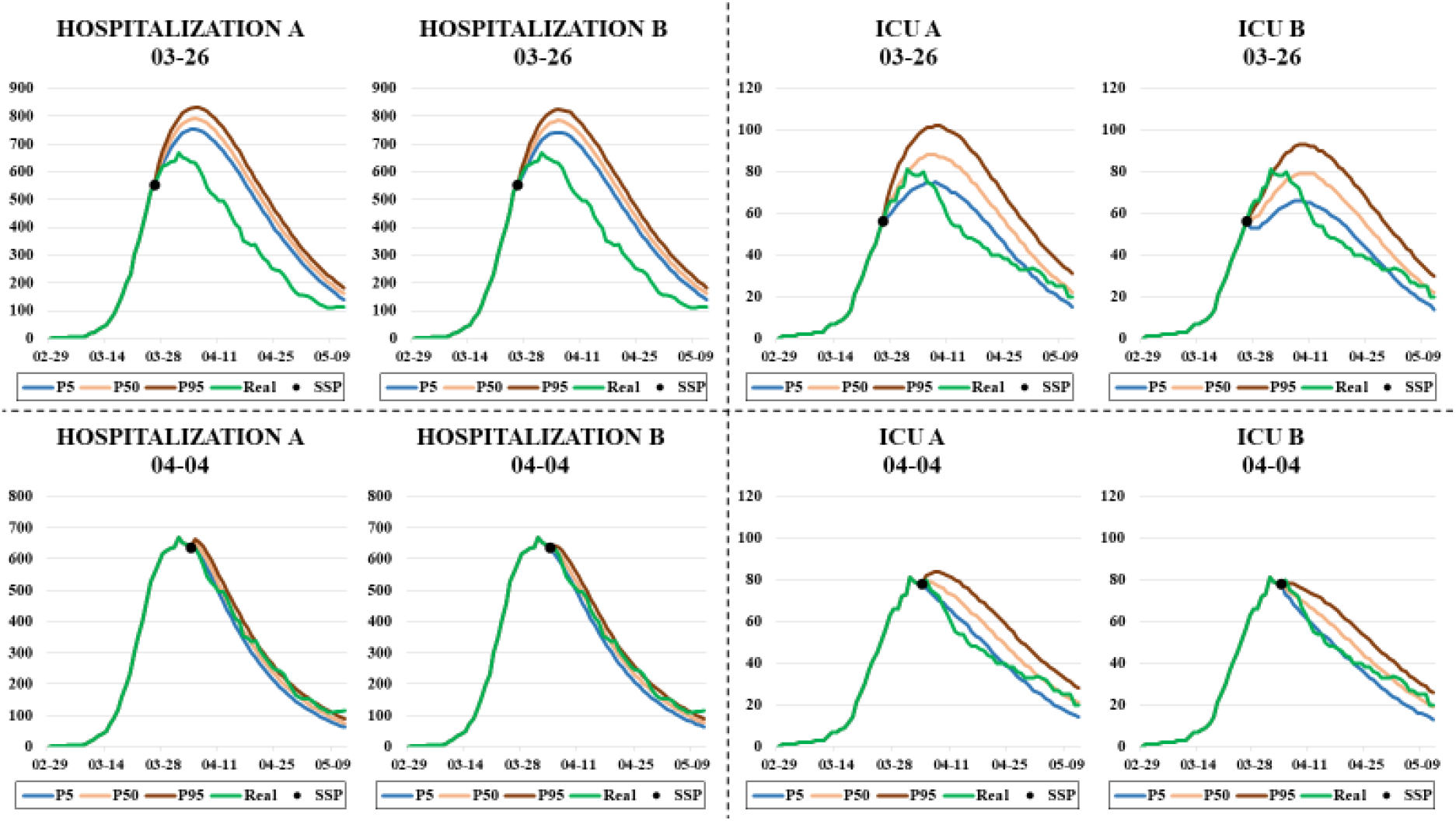
Comparative graphs on the predictions made with different levels of information (scenarios A and B). Each graph shows the predictions made for the number of beds occupied in the hospitals and the ICUs of Navarre and the real occupancies. The four predictions at the top side were made on March 26, while those four at the low were made on April 4.

In view of the results, it can be seen that there is no great difference between the two types of simulation, which is very encouraging since the usefulness of the simulator is demonstrated even when all the disaggregated information on patients is not available. In addition to this, it is observed that the results are more accurate on the second date because more information is available and it is somewhat easier to predict the occupation of beds once the peak occupancy has been exceeded.

## 8 Conclusions

Healthcare systems are overburdened due to a large demand for healthcare services from COVID-19 patients that leads to strained ICU’s capacity and overworked healthcare workers. Having accurate predictions of the resources needed for taking care of these patients are essential for planning in advance the necessary resources and reduce the pressure on the system and stress on the healthcare staff.

Normally, managers can create contingency plans to deploy sufficient resources to meet the increase in demand based on the predictions made about one week in advance. In this paper, we have developed a DES model for predicting the need for hospital resources, particularly ward and ICU beds. The simulation model is fed by the predictions of new hospitalizations made, directly or indirectly, through a PG model. The Gompertz growth model has been selected after analyzing the fitting and forecasting properties of four PG models: logistic, Richards, Stannard, and Gompertz. An ensemble of these models could improve predictions, but this research is out of the scope of this paper and it is left for future work. Besides, other factors influencing the consumption of resources are the age and the Adjusted Morbidity Group (AMG) whose inclusion in the stochastic models for the length of stay can provide more accurate predictions.

The structural simplicity of the simulation model makes it appropriate for general use, i.e., it can be adapted to estimate the bed needs in any geographic area. The growth model is simple enough to fit it to available data or, in the absence of it, to be estimated by educated guesses of experts.

It is worthy to mention the strength of simulation models in this context of uncertainty: their capability to run what-if scenarios that allow decision-makers to explore the consequences of different policy choices, like the location and number of additional healthcare resources needed for COVID-19 patients given the uncertainty in demand. The simulation model is data-driven, patients arrivals and length of stays can be estimated from data, but it has also the flexibility of allowing the simulation from the input determined by the user to explore additional scenarios.

From a technical and methodological point of view, a distinct feature of the simulation model is its focus on the transition period of the health system instead of the stationary state as it is usual in the simulation studies or transition periods but after regeneration points. This transition period reproduced by the simulator is unique as the outbreak evolves with no regeneration points. Therefore, the accurate representation of the initial health system state plays an important role. The simulation of the remaining length of stay of each patient already admitted in the hospital has shown to be a key point to project smoothly the dynamics of the health system and linking it (and mixing it) with the new dynamics obtained from the simulation of the arrivals and stays of the new incoming patients. However, the simulation of the remaining length of stay depends on the information level about the admitted patients. In this paper, we have considered four different levels of information that grades from the complete information to the patient level (knowing exactly the dates of admission and discharge) to the only knowledge of the number of admitted patients. The simulation model can work with any of these information levels, making it a tool of very general application.

This simulation paradigm is most suitable for the realistic representation of processes in health services, which makes it more credible and easier to understand by the managers that will have to rely on their results to make their decisions. The involvement in the development of the simulation model of the person in charge of the hospital system logistic person has been crucial to obtaining a user-centered simulator, which was daily feed with the new data gathered by the hospital administrative information system.

## Data Availability

Historical data is available on publicly accessible web pages. The patient data used in the case study is not authorized to be made public.

https://coronavirus.jhu.edu/map.html

https://ec.europa.eu/info/live-work-travel-eu/health/coronavirus-response_en

https://www.who.int/emergencies/diseases/novel-coronavirus-2019

https://www.worldometers.info/coronavirus/

## Declarations

### Funding

the grant MTM2016-77015-R (AEI, FEDER EU).

### Conflicts of interest/Competing interests

Declarations of interest: none.

Availability of data and material:

### Code availability

custom code programming with Python.

### Authors’ contributions

**Daniel Garcia-Vicuña:** Methodology, Programming, Formal Analysis, Investigation, Data Curation, Writing. **Laida Esparza:** Conceptualization, Validation, Resources, Investigation, Data Curation, Writing. **Fermin Mallor:** Conceptualization, Methodology, Formal Analysis, Data Curation, Writing, Supervision, Funding Acquisition.

## Acknowledgments

This paper has been supported by grant MTM2016-77015-R (AEI, FEDER EU). The authors are grateful to the medical staff of the main hospital in Navarre for providing data and validating the simulation model.

## APPENDICES

### A. Results of positive cases predictions for the following 5, 10, and 15 days, at 25%, 45%, and 65% of total cases detected

**Table 5.** *MAE calculated for the fit of each model at 25% of total cases detected*.

| # | Country | Date | Total positive cases | Logistic | Gompertz | Richards | Stannard |
| --- | --- | --- | --- | --- | --- | --- | --- |
| 1 | USA | 2020-04-12 | 529,951 | 2,233.0 | 554.9 | **553.7** | **553.7** |
| 2 | Brazil | 2020-05-16 | 218,223 | 1,497.5 | 1,099.9 | **1,098.4** | 1,100.4 |
| 3 | Russia | 2020-05-04 | 134,687 | 564.2 | 381.8 | **357.2** | **357.2** |
| 4 | India | 2020-05-16 | 85,940 | 601.4 | **350.0** | **350.0** | **350.1** |
| 5 | UK | 2020-04-12 | 78,991 | 320.6 | 249.7 | **209.7** | **209.7** |
| 6 | Spain | 2020-03-26 | 66,460 | 297.1 | **147.8** | **147.8** | **147.9** |
| 7 | Italy | 2020-03-24 | 63,927 | 269.7 | **150.8** | 156.7 | 156.7 |
| 8 | Peru | 2020-05-08 | 58,526 | 608.4 | **385.0** | **385.1** | **385.1** |
| 9 | Iran | 2020-04-02 | 47,593 | 1,192.5 | **969.6** | **969.6** | **969.7** |
| 10 | Germany | 2020-03-28 | 48,582 | **269.9** | 315.8 | 287.4 | 287.4 |
| 11 | Turkey | 2020-04-11 | 47,029 | 639.9 | **272.4** | **272.5** | **272.6** |
| 12 | Chile | 2020-05-18 | 43,781 | 741.7 | **732.2** | 735.8 | 735.8 |
| 13 | France | 2020-03-30 | 40,174 | 163.2 | **92.6** | **93.0** | 94.6 |
| 14 | Mexico | 2020-05-06 | 40,186 | 207.8 | 130.5 | **128.3** | **128.3** |
| 15 | Pakistan | 2020-05-15 | 37,218 | 304.3 | **223.5** | **223.1** | 223.5 |
| 16 | Saudi Arabia | 2020-05-07 | 31,938 | **194.3** | 281.4 | **195.2** | **195.2** |
| 17 | Canada | 2020-04-14 | 25,663 | 97.0 | 83.7 | **57.3** | **57.3** |
| 18 | Bangladesh | 2020-05-18 | 22,268 | 270.9 | **134.5** | **134.5** | **134.5** |
| 19 | China | 2020-02-05 | 24,320 | 164.3 | **114.1** | **114.1** | **114.1** |
| 20 | Qatar | 2020-05-09 | 20,201 | 147.6 | 206.0 | **139.2** | **139.2** |

**Table 6.**
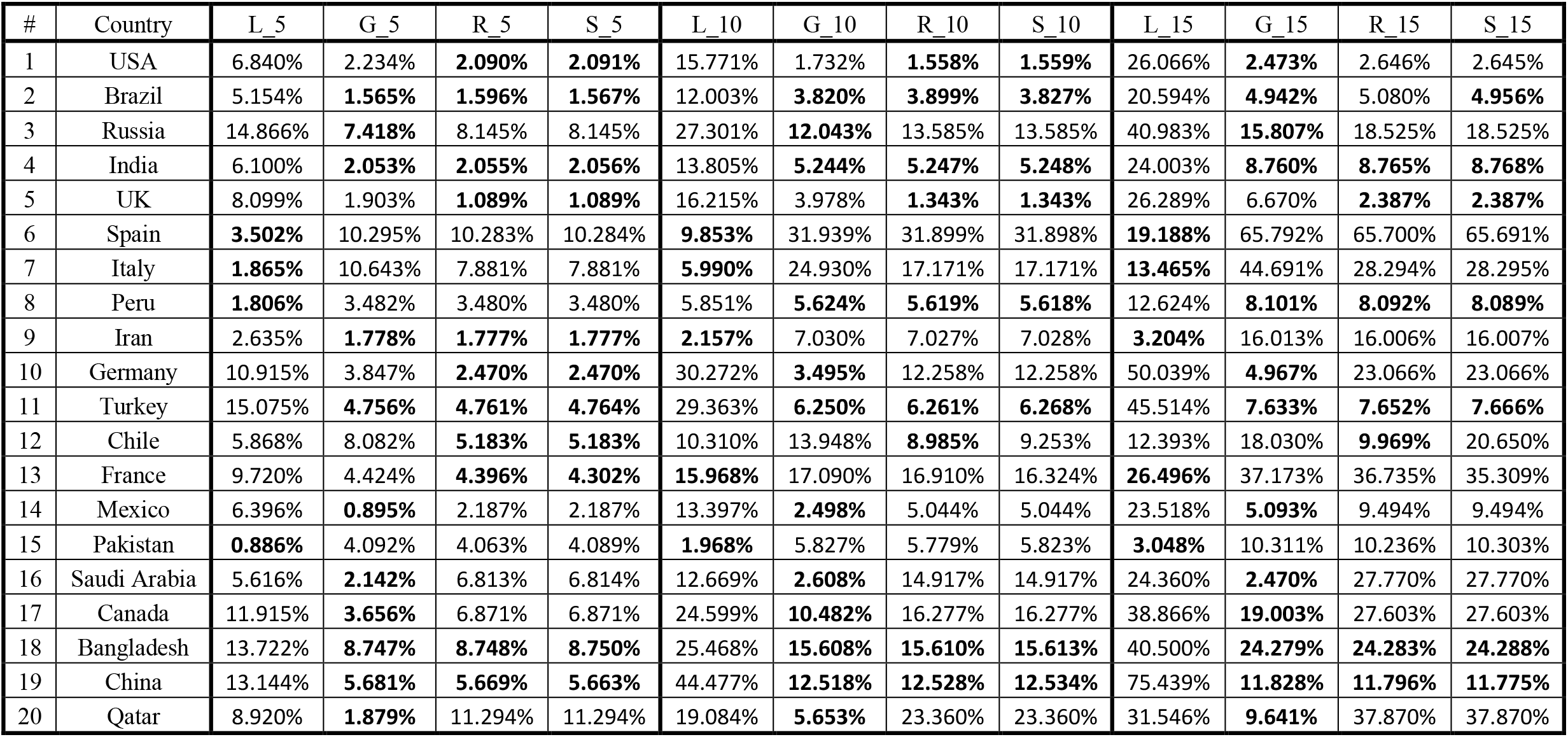
*Normalized MAEs obtained for each prediction and model at 25% of total cases detected*.

| # | Country | L 5 | G 5 | R 5 | S 5 | L 10 | G 10 | R 10 | S 10 | L 15 | G 15 | R 15 | S 15 |
| --- | --- | --- | --- | --- | --- | --- | --- | --- | --- | --- | --- | --- | --- |
| 1 | USA | 6.840% | 2.234% | **2.090%** | **2.091%** | 15.771% | 1.732% | **1.558%** | **1.559%** | 26.066% | **2.473%** | 2.646% | 2.645% |
| 2 | Brazil | 5.154% | **1.565%** | **1.596%** | **1.567%** | 12.003% | **3.820%** | **3.899%** | **3.827%** | 20.594% | **4.942%** | 5.080% | **4.956%** |
| 3 | Russia | 14.866% | **7.418%** | 8.145% | 8.145% | 27.301% | **12.043%** | 13.585% | 13.585% | 40.983% | **15.807%** | 18.525% | 18.525% |
| 4 | India | 6.100% | **2.053%** | **2.055%** | **2.056%** | 13.805% | **5.244%** | **5.247%** | **5.248%** | 24.003% | **8.760%** | **8.765%** | **8.768%** |
| 5 | UK | 8.099% | 1.903% | **1.089%** | **1.089%** | 16.215% | 3.978% | **1.343%** | **1.343%** | 26.289% | 6.670% | **2.387%** | **2.387%** |
| 6 | Spain | **3.502%** | 10.295% | 10.283% | 10.284% | **9.853%** | 31.939% | 31.899% | 31.898% | **19.188%** | 65.792% | 65.700% | 65.691% |
| 7 | Italy | **1.865%** | 10.643% | 7.881% | 7.881% | **5.990%** | 24.930% | 17.171% | 17.171% | **13.465%** | 44.691% | 28.294% | 28.295% |
| 8 | Peru | **1.806%** | 3.482% | 3.480% | 3.480% | 5.851% | **5.624%** | **5.619%** | **5.618%** | 12.624% | **8.101%** | **8.092%** | **8.089%** |
| 9 | Iran | 2.635% | **1.778%** | **1.777%** | **1.777%** | **2.157%** | 7.030% | 7.027% | 7.028% | **3.204%** | 16.013% | 16.006% | 16.007% |
| 10 | Germany | 10.915% | 3.847% | **2.470%** | **2.470%** | 30.272% | **3.495%** | 12.258% | 12.258% | 50.039% | **4.967%** | 23.066% | 23.066% |
| 11 | Turkey | 15.075% | **4.756%** | **4.761%** | **4.764%** | 29.363% | **6.250%** | **6.261%** | **6.268%** | 45.514% | **7.633%** | **7.652%** | **7.666%** |
| 12 | Chile | 5.868% | 8.082% | **5.183%** | **5.183%** | 10.310% | 13.948% | **8.985%** | 9.253% | 12.393% | 18.030% | **9.969%** | 20.650% |
| 13 | France | 9.720% | 4.424% | **4.396%** | **4.302%** | **15.968%** | 17.090% | 16.910% | 16.324% | **26.496%** | 37.173% | 36.735% | 35.309% |
| 14 | Mexico | 6.396% | **0.895%** | 2.187% | 2.187% | 13.397% | **2.498%** | 5.044% | 5.044% | 23.518% | **5.093%** | 9.494% | 9.494% |
| 15 | Pakistan | **0.886%** | 4.092% | 4.063% | 4.089% | **1.968%** | 5.827% | 5.779% | 5.823% | **3.048%** | 10.311% | 10.236% | 10.303% |
| 16 | Saudi Arabia | 5.616% | **2.142%** | 6.813% | 6.814% | 12.669% | **2.608%** | 14.917% | 14.917% | 24.360% | **2.470%** | 27.770% | 27.770% |
| 17 | Canada | 11.915% | **3.656%** | 6.871% | 6.871% | 24.599% | **10.482%** | 16.277% | 16.277% | 38.866% | **19.003%** | 27.603% | 27.603% |
| 18 | Bangladesh | 13.722% | **8.747%** | **8.748%** | **8.750%** | 25.468% | **15.608%** | **15.610%** | **15.613%** | 40.500% | **24.279%** | **24.283%** | **24.288%** |
| 19 | China | 13.144% | **5.681%** | **5.669%** | **5.663%** | 44.477% | **12.518%** | **12.528%** | **12.534%** | 75.439% | **11.828%** | **11.796%** | **11.775%** |
| 20 | Qatar | 8.920% | **1.879%** | 11.294% | 11.294% | 19.084% | **5.653%** | 23.360% | 23.360% | 31.546% | **9.641%** | 37.870% | 37.870% |

**Table 7.** *MAE calculated for the fit of each model at 40% of total cases detected*.

| # | Country | Date | Total positive cases | Logistic | Gompertz | Richards | Stannard |
| --- | --- | --- | --- | --- | --- | --- | --- |
| 1 | USA | 2020-04-23 | 842,629 | 6,372.8 | **1,421.9** | **1,422.8** | 1,433.3 |
| 2 | Brazil | 2020-05-24 | 347,398 | 2,085.6 | 1,422.1 | **1,413.6** | 1,422.7 |
| 3 | Russia | 2020-05-12 | 221,344 | 1,316.9 | **521.2** | **521.3** | **521.5** |
| 4 | India | 2020-05-25 | 138,845 | 882.2 | **480.3** | **480.4** | **480.5** |
| 5 | UK | 2020-04-20 | 120,067 | 617.4 | 320.3 | **257.3** | **257.3** |
| 6 | Spain | 2020-03-31 | 104,267 | 351.9 | 438.7 | **343.9** | **343.9** |
| 7 | Italy | 2020-03-30 | 97,689 | 374.7 | 343.5 | **205.8** | **205.8** |
| 8 | Peru | 2020-05-18 | 92,273 | 759.3 | 483.2 | **482.1** | **482.1** |
| 9 | Iran | 2020-04-16 | 76,389 | 1,071.3 | 1,201.7 | **1,043.0** | **1,043.0** |
| 10 | Germany | 2020-04-03 | 79,696 | 486.0 | 339.2 | **338.2** | **338.2** |
| 11 | Turkey | 2020-04-17 | 74,193 | 795.4 | **345.4** | **345.5** | **345.7** |
| 12 | Chile | 2020-05-26 | 73,997 | **854.4** | 858.2 | **854.4** | **854.4** |
| 13 | France | 2020-04-04 | 64,338 | 260.8 | **200.4** | 219.2 | 219.2 |
| 14 | Mexico | 2020-05-15 | 62,527 | 438.7 | **151.8** | **151.8** | **151.9** |
| 15 | Pakistan | 2020-05-27 | 59,151 | 335.9 | **276.6** | 285.4 | 285.4 |
| 16 | Saudi Arabia | 2020-05-17 | 52,016 | 360.6 | 273.4 | **256.6** | **256.6** |
| 17 | Canada | 2020-04-23 | 40,179 | 405.1 | **175.7** | **175.7** | 175.7 |
| 18 | Bangladesh | 2020-05-26 | 35,585 | 434.7 | **270.4** | **270.4** | 270.5 |
| 19 | China | 2020-02-08 | 34,625 | 185.9 | 130.2 | **126.6** | **126.6** |
| 20 | Qatar | 2020-05-18 | 32,604 | 274.1 | **216.8** | **216.8** | **216.8** |

**Table 8.** *Normalized MAEs obtained for each prediction and model at 40% of total cases detected*.

| # | Country | L 5 | G 5 | R 5 | S 5 | L 10 | G 10 | R 10 | S 10 | L 15 | G 15 | R 15 | S 15 |
| --- | --- | --- | --- | --- | --- | --- | --- | --- | --- | --- | --- | --- | --- |
| 1 | USA | 9.702% | **3.501%** | **3.503%** | **3.525%** | 15.921% | **6.527%** | **6.531%** | **6.567%** | 22.590% | **10.112%** | **10.117%** | **10.167%** |
| 2 | Brazil | **1.165%** | 3.163% | 2.984% | 3.161% | **2.858%** | 3.924% | 3.617% | 3.919% | 6.209% | 6.041% | **5.547%** | 6.032% |
| 3 | Russia | 3.420% | **2.303%** | **2.301%** | **2.299%** | **5.698%** | 6.093% | 6.090% | 6.085% | **9.245%** | 10.728% | 10.721% | 10.714% |
| 4 | India | 4.669% | **1.331%** | **1.334%** | **1.333%** | 9.212% | **2.274%** | **2.278%** | **2.277%** | 16.059% | **3.695%** | **3.701%** | **3.700%** |
| 5 | UK | 6.330% | 1.005% | **0.710%** | **0.711%** | 11.831% | **0.913%** | 2.210% | 2.210% | 19.098% | **1.746%** | 5.344% | 5.344% |
| 6 | Spain | 6.632% | **5.630%** | 5.905% | 5.906% | 13.015% | 11.927% | **11.725%** | **11.725%** | 20.529% | **18.121%** | 18.794% | 18.795% |
| 7 | Italy | 3.211% | 6.294% | **1.096%** | **1.096%** | 8.347% | 9.938% | **1.571%** | **1.571%** | 14.945% | 13.137% | **4.334%** | **4.334%** |
| 8 | Peru | 6.317% | **0.817%** | 1.035% | 1.035% | 11.975% | **1.669%** | 2.145% | 2.146% | 22.254% | **6.073%** | 6.872% | 6.873% |
| 9 | Iran | **0.582%** | 6.097% | 1.628% | 1.628% | **1.757%** | 8.452% | 3.338% | 3.337% | **3.372%** | 10.640% | 5.413% | 5.413% |
| 10 | Germany | 11.428% | **2.083%** | 2.224% | 2.220% | 19.849% | **2.095%** | 2.390% | 2.381% | 27.383% | **1.755%** | **1.802%** | **1.801%** |
| 11 | Turkey | 7.319% | **1.436%** | **1.432%** | **1.429%** | 13.559% | **3.472%** | **3.463%** | **3.457%** | 19.637% | **6.500%** | **6.486%** | **6.476%** |
| 12 | Chile | **1.645%** | **1.636%** | **1.645%** | **1.645%** | 5.823% | **5.639%** | 5.823% | 5.817% | 14.435% | 14.059% | 14.432% | **7.991%** |
| 13 | France | **2.158%** | 12.580% | 6.941% | 6.940% | **3.202%** | 22.970% | 9.453% | 9.451% | **7.545%** | 35.881% | 10.903% | 10.900% |
| 14 | Mexico | 7.023% | **2.499%** | **2.501%** | **2.503%** | 12.126% | **3.558%** | **3.563%** | **3.566%** | 19.255% | **5.141%** | **5.149%** | **5.154%** |
| 15 | Pakistan | 3.195% | 1.697% | **1.489%** | **1.489%** | 11.675% | **4.830%** | 7.837% | 7.837% | 23.909% | **11.558%** | 17.595% | 17.595% |
| 16 | Saudi Arabia | 9.921% | **4.418%** | 5.822% | 5.822% | 15.880% | **5.955%** | 8.499% | 8.499% | 20.963% | **5.429%** | 9.512% | 9.512% |
| 17 | Canada | 12.339% | **6.475%** | **6.477%** | **6.478%** | 19.244% | **9.745%** | **9.749%** | **9.751%** | 27.788% | **14.521%** | **14.526%** | **14.529%** |
| 18 | Bangladesh | 4.289% | **1.024%** | **1.025%** | **1.025%** | 11.261% | **3.951%** | **3.955%** | **3.954%** | 20.791% | **7.544%** | **7.550%** | **7.550%** |
| 19 | China | 11.511% | 10.730% | **10.225%** | **10.225%** | 37.831% | **12.943%** | 16.756% | 16.756% | 52.069% | **10.778%** | 15.505% | 15.506% |
| 20 | Qatar | 5.565% | **1.188%** | **1.204%** | **1.211%** | 9.960% | **1.348%** | **1.379%** | **1.392%** | 16.349% | **1.774%** | **1.827%** | **1.849%** |

**Table 9.** *MAE calculated for the fit of each model at 65% of total cases detected*.

| # | Country | Date | Total positive cases | Logistic | Gompertz | Richards | Stannard |
| --- | --- | --- | --- | --- | --- | --- | --- |
| 1 | USA | 2020-04-13 | 1,369,964 | 20,036.0 | **7,343.8** | 7,345.7 | 7,346.8 |
| 2 | Brazil | 2020-05-31 | 584,016 | 2,654.8 | **1,857.5** | 1,908.6 | 1,908.4 |
| 3 | Russia | 2020-05-19 | 344,481 | 1,469.0 | 1,097.0 | **846.3** | **846.3** |
| 4 | India | 2020-04-13 | 216,919 | 1,113.0 | **511.4** | **511.6** | **511.7** |
| 5 | UK | 2020-04-30 | 194,990 | 2,136.8 | **593.2** | **593.2** | **593.3** |
| 6 | Spain | 2020-06-04 | 163,472 | 917.7 | 639.9 | **382.2** | **382.2** |
| 7 | Italy | 2020-04-15 | 156,363 | 1,420.8 | 585.3 | **526.1** | **526.1** |
| 8 | Peru | 2020-05-27 | 155,671 | 1,757.1 | **792.6** | **792.9** | **793.1** |
| 9 | Iran | 2020-06-06 | 122,492 | 3,124.1 | **1,912.6** | **1,912.7** | **1,912.8** |
| 10 | Germany | 2020-05-31 | 123,016 | 907.9 | 419.2 | **400.3** | **400.3** |
| 11 | Turkey | 2020-05-08 | 117,589 | 1,222.6 | 528.2 | **463.3** | **463.3** |
| 12 | Chile | 2020-06-05 | 113,628 | 1,011.6 | 1,038.2 | **844.2** | **844.2** |
| 13 | France | 2020-02-13 | 103,573 | 495.0 | 525.4 | **350.4** | **350.4** |
| 14 | Mexico | 2020-05-30 | 101,238 | 746.9 | **237.1** | **237.2** | **237.3** |
| 15 | Pakistan | 2020-04-13 | 93,983 | 918.8 | **565.8** | **565.8** | **566.0** |
| 16 | Saudi Arabia | 2020-05-31 | 83,384 | 683.4 | 453.3 | **430.7** | **430.7** |
| 17 | Canada | 2020-05-19 | 64,922 | 836.0 | **346.9** | **346.9** | **347.0** |
| 18 | Bangladesh | 2020-04-13 | 57,563 | 534.4 | **310.3** | **310.4** | **310.4** |
| 19 | China | 2020-04-30 | 59,865 | 841.5 | **556.6** | **556.6** | **556.7** |
| 20 | Qatar | 2020-06-04 | 52,907 | 352.0 | 199.7 | **199.6** | **199.6** |

**Table 10.** *Normalized MAEs obtained for each prediction and model at 65% of total cases detected*.

| # | Country | L 5 | G 5 | R 5 | S 5 | L 10 | G 10 | R 10 | S 10 | L 15 | G 15 | R 15 | S 15 |
| --- | --- | --- | --- | --- | --- | --- | --- | --- | --- | --- | --- | --- | --- |
| 1 | USA | 7.840% | **3.355%** | **3.356%** | **3.356%** | 10.822% | **4.910%** | **4.911%** | **4.911%** | 13.993% | **6.696%** | **6.698%** | **6.698%** |
| 2 | Brazil* | 2.720% | **1.445%** | **1.396%** | **1.397%** | 3.898% | 3.799% | **3.153%** | **3.156%** | 4.007% | 4.631% | **3.863%** | **3.867%** |
| 3 | Russia | 4.285% | 1.705% | **0.901%** | **0.902%** | 7.670% | **1.598%** | 2.524% | 2.524% | 11.790% | **1.254%** | 4.822% | 4.822% |
| 4 | India* | 4.293% | **0.859%** | **0.861%** | **0.861%** | 7.276% | **0.607%** | **0.609%** | **0.610%** | 8.062% | **0.575%** | **0.577%** | **0.578%** |
| 5 | UK | 9.641% | **4.441%** | **4.443%** | **4.444%** | 12.914% | **5.616%** | **5.618%** | **5.620%** | 16.050% | **6.681%** | **6.684%** | **6.687%** |
| 6 | Spain | 6.365% | **0.354%** | 2.477% | 2.477% | 9.902% | **0.485%** | 4.430% | 4.430% | 13.199% | **1.001%** | 6.451% | 6.451% |
| 7 | Italy | 7.789% | **2.131%** | 3.037% | 3.037% | 11.523% | **3.678%** | 4.993% | 4.993% | 15.241% | **5.390%** | 7.116% | 7.116% |
| 8 | Peru | 10.445% | **6.119%** | **6.121%** | **6.122%** | 12.525% | **5.208%** | **5.210%** | **5.212%** | 15.847% | **4.811%** | **4.815%** | **4.818%** |
| 9 | Iran | 14.856% | **10.700%** | **10.702%** | **10.702%** | 18.765% | **13.960%** | **13.962%** | **13.963%** | 23.212% | **17.807%** | **17.809%** | **17.810%** |
| 10 | Germany | 4.053% | 2.415% | **1.864%** | **1.864%** | 7.360% | 2.354% | **1.473%** | **1.473%** | 10.611% | 2.080% | **1.118%** | **1.118%** |
| 11 | Turkey | 4.858% | 1.079% | **0.484%** | **0.484%** | 7.419% | **0.972%** | 1.333% | 1.333% | 10.097% | **0.817%** | 2.504% | 2.504% |
| 12 | Chile* | **6.750%** | 9.179% | 7.516% | 7.517% | **11.535%** | 16.538% | 17.398% | 17.400% | **12.420%** | 18.133% | 19.826% | 19.827% |
| 13 | France | 5.234% | **1.690%** | 1.813% | 1.813% | 7.530% | 2.840% | **2.534%** | **2.534%** | 9.905% | 3.784% | **3.530%** | **3.530%** |
| 14 | Mexico | 5.207% | **1.082%** | **1.084%** | **1.085%** | 8.908% | **1.584%** | **1.587%** | **1.588%** | 11.785% | **2.536%** | **2.537%** | **2.538%** |
| 15 | Pakistan** | 11.997% | **9.348%** | **9.351%** | **9.351%** | 18.651% | **14.243%** | **14.246%** | **14.247%** | - | - | - | - |
| 16 | Saudi Arabia | **0.598%** | 4.724% | 3.395% | 3.395% | 4.029% | 4.289% | **2.612%** | **2.612%** | 10.167% | **3.939%** | 4.547% | 4.547% |
| 17 | Canada | 5.661% | **0.861%** | **0.862%** | **0.863%** | 7.801% | **0.674%** | **0.676%** | **0.677%** | 10.379% | **0.791%** | **0.793%** | **0.794%** |
| 18 | Bangladesh | 5.516% | **2.097%** | **2.099%** | **2.099%** | 9.975% | **3.061%** | **3.064%** | **3.065%** | - | - | - | - |
| 19 | China | 14.044% | **5.492%** | **5.495%** | **5.495%** | 14.165% | **8.987%** | **8.976%** | **8.973%** | **14.158%** | 16.806% | 16.781% | 16.776% |
| 20 | Qatar | 4.045% | **0.780%** | **0.777%** | **0.775%** | 5.683% | **2.302%** | **2.293%** | **2.286%** | 7.766% | **4.616%** | **4.600%** | **4.589%** |

## Notes

### Competing Interest Statement

The authors have declared no competing interest.

### Author Declarations

Public University of Navarre

